# Stratifying elicited antibody dynamics after two doses of SARS-CoV-2 vaccine in a community-based cohort in Fukushima, Japan

**DOI:** 10.1101/2022.06.11.22276266

**Authors:** Naotoshi Nakamura, Yurie Kobashi, Kwang Su Kim, Yuta Tani, Yuzo Shimazu, Tianchen Zhao, Yoshitaka Nishikawa, Fumiya Omata, Moe Kawashima, Makoto Yoshida, Toshiki Abe, Yoshika Saito, Yuki Senoo, Saori Nonaka, Morihito Takita, Chika Yamamoto, Takeshi Kawamura, Akira Sugiyama, Aya Nakayama, Yudai Kaneko, Hyeongki Park, Yong Dam Jeong, Daiki Tatematsu, Marwa Akao, Yoshitaka Sato, Shoya Iwanami, Yasuhisa Fujita, Masatoshi Wakui, Kazuyuki Aihara, Tatsuhiko Kodama, Kenji Shibuya, Shingo Iwami, Masaharu Tsubokura

## Abstract

Recent studies have provided insights into the effect of vaccine boosters on recall immunity. Given the limited global supply of COVID-19 vaccines, identifying vulnerable populations with lower sustained vaccine-elicited antibody titers is important for targeting individuals for booster vaccinations. Here we investigated longitudinal data in a cohort of 2,526 people in Fukushima, Japan, from April 2021 to December 2021. Antibody titers following two doses of a COVID-19 vaccine were repeatedly monitored and information on lifestyle habits, comorbidities, adverse reactions, and medication use was collected. Using mathematical modeling and machine learning, we stratified the time-course patterns of antibody titers and identified vulnerable populations with low sustained antibody titers. Moreover, we showed that only 5.7% of the participants in our cohort were part of the “durable” population with high sustained antibody titers, which suggests that this durable population might be overlooked in small cohorts. We also found large variation in antibody waning within our cohort. There is a potential usefulness of our approach for identifying the neglected vulnerable population.

## Main Text

Primary two-dose coronavirus disease 2019 (COVID-19) vaccination leads to rapid immunity, providing protection against severe acute respiratory syndrome coronavirus 2 (SARS-CoV-2) infection^1^. Although COVID-19 vaccines still offer good protection against severe disease and death for months after the second dose owing to the durability of immunity^2–4^, the waning of vaccine efficacy has become a major concern in the era of “living with COVID-19”^5–9^. The rapid decline in vaccine-elicited antibodies results in breakthrough infections and adverse health outcomes caused by emerging variants of concern (VOCs), including the Beta, Delta, and Omicron variants^10–13^. Additional third and fourth doses of vaccine induce a high-level antibody response against VOCs^13–17^, implying a need for additional booster vaccinations as a future pandemic exit strategy^18–20^. Few studies, however, have quantitatively stratified and assessed the variation in waning of antibody titers at both the population and individual level in a community-based large cohort.

At the population level, it is well known that the older a person is, the faster their protection weakens after the second vaccination^7, 8^, and that people with underlying medical conditions tend to induce an insufficient antibody response^8, 21^. These populations are generally the highest priority group for booster vaccinations. There is, however, large variation in the vaccine-elicited antibody response at the individual level, even within the same age group^6–8, 15, 21, 22^. Assessing the factors that affect such variations will shed further light on our understanding of waning of protection and identify potentially vulnerable populations that may be a priority for booster vaccination^8, 21^. In the long-run, personalized vaccination will be required, because continuous vaccinations after the third dose to the whole population may be neither sustainable nor feasible globally. One approach to assess variation within the same group of people is the stratified analysis of time-dependent vaccine-elicited antibody dynamics.

Although the mean time-dependent vaccine efficacy for symptomatic and severe SARS-CoV-2 infections has been assessed through cross-sectional and longitudinal studies at the population level^2, 3^, many relevant aspects of the individual-level time course of antibody dynamics and the variations in individual-level dynamics are not well understood. In this study, we collected longitudinal datasets on individual antibody responses, together with personal information including lifestyle habits, comorbidities, adverse reactions, and medication use, from more than 2,526 individuals of a community-based cohort in Fukushima, Japan. Taking advantage of this unique cohort and using mathematical modeling and machine-learning approaches, we stratified the elicited immune response after two doses of a COVID-19 vaccine by reconstructing individual antibody dynamics. Furthermore, we explored the factors associated with the durability of vaccine response. Our aim was to understand individual-level variation in vaccine-elicited immune responses at the time of booster vaccinations and to inform long-term COVID-19 vaccination strategies.

## Results

### The Fukushima vaccination cohort

This study was conducted from April 2021 to December 2021 in Fukushima, Japan. Our vaccination cohort consisted of participants from a primarily rural area where COVID-19 prevalence was relatively low: Soma City, Minami Soma City, and Hirata village (**Fig 1A**). The participants included health care workers, frontline workers, administrative officers, general residents, and residents of long-term care facilities. In total, 2,526 participants who had been vaccinated with the Pfizer BNT162b2 or Moderna mRNA-1273 vaccine were recruited, and 2,159 participants were included in the final data analysis, respectively (see **Fig 1B** and **Methods** in details). The age and sex distributions of the participants are shown in **Fig 1C**, and the sample characteristics and information on adverse vaccine events stratified by age are provided in **Extended Data Table 1**. A portion of this cohort was described previously for the time period extending to 6 months after the first dose of mRNA vaccine^23, 24^.

**Figure 1.**
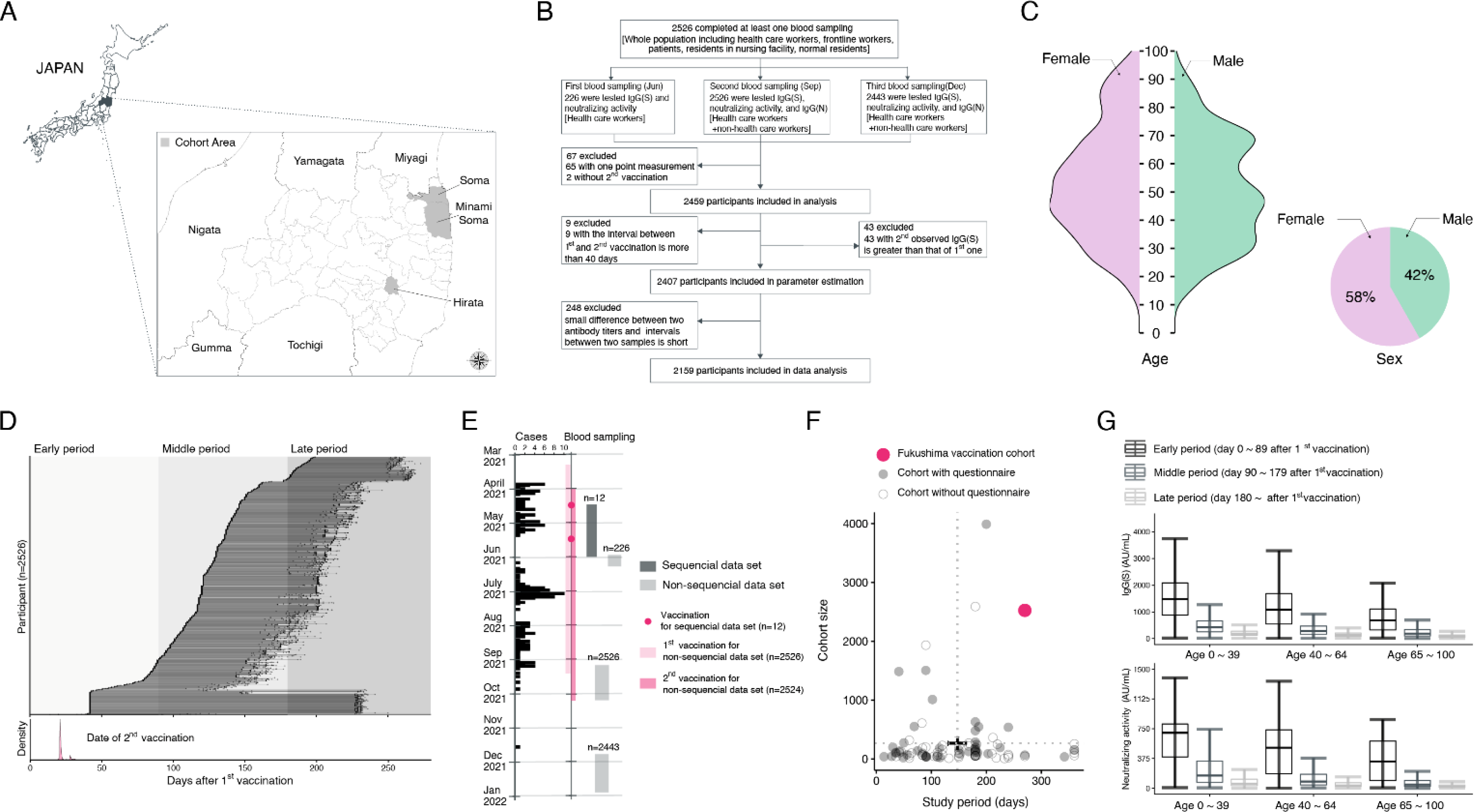
Characteristics of the Fukushima vaccination cohort. **(A)** Locations of Soma city, Minami Soma city, and Hirata village in Fukushima prefecture are described. **(B)** Flowchart of the vaccination cohort along with the number of participants, testing, and inclusion criteria for our analysis are described. **(C)** Age and sex distributions in the cohort are shown. **(D)** Timeline of sample collection for each cohort participant (N=2,526 participants, 5195 total samples) is described. The timings of blood samplings are indicated in black circles. Shaded areas indicate early (<89 days), middle (90-179 days), and late (>180 days) time periods after the first vaccination. Dates for the second vaccination are shown as the distributions in the bottom panel. **(E)** Vaccination and blood sampling periods in the cohort along with the number of COVID-19 cases (i.e., cases) in Soma city, Minami Soma city, and Hirata village^48, 49^ are shown. **(F)** Distribution of the study periods and the number of participants (i.e., cohort size) of previously reported cohorts extracted by literature review (**Supplementary Note 1**) are plotted along with the Fukushima vaccination cohort (the red mark). The closed and open marks correspond to cohorts with and without questionnaire data. The origin of coordinates corresponds to the median of the period and number of the cohorts, and the whiskers indicate corresponding standard derivations. **(G)** Longitudinal IgG(S) and neutralizing activity measured by CLIA are separately plotted by time after the first vaccination and age.

Here we investigated antibody titers in the Fukushima cohort in individuals sampled longitudinally (serum was collected at 2 or 3 different timepoints) for around 4 to 9 months after the second primary dose of mRNA vaccine (see **Fig 1DE** for details). Notably, the number of SARS-CoV-2 infections in this rural area was extremely low (**Fig 1E**), so that we could minimize the influence of breakthrough natural infections. The Fukushima cohort, which is large enough and with a long study period (**Fig 1F**), is an ideal longitudinal cohort for quantifying and stratifying vaccine-elicited antibody dynamics (**Supplementary Note 1** and **Supplementary Table 1**). In particular, compared with the largest cohort of more than 3,900 participants from Israel,^6^ the Fukushima cohort is community-based, includes non-health workers, has very few dropouts among more than 2,000 identical individuals who were consecutively sampled (only 3.3%), includes all necessary information for all participants, and includes measures of several modalities of antibody titers including neutralizing activity (**Supplementary Table 2**).

We performed chemiluminescent immunoassay (CLIA) to measure antibody titers as a measure of humoral immune status after the first COVID-19 vaccination (i.e., a total of 5195 IgG(S), 5195 neutralization activity, and 4969 IgG(N) assays were performed) (**Methods**). **Fig 1G** shows the overall profile of IgG(S) and neutralization activity in this study. We investigated longitudinal data for IgG(S) in the same individuals because IgG(S) covers a wider range of antibody responses and is more sensitive than neutralization activity. In fact, these two measurements are highly correlated with each other (correlation coefficient of 0.93) (**Extended Data Fig 1AB**), and previous studies showed that neutralizing antibody and IgG(S) titers correlate with vaccine-mediated protection, even against VOCs (i.e., vaccine efficacy)^1–3^. This is because vaccines containing the original Wuhan virus spike protein induce variant-reactive memory B cells targeting multiple different VOCs, including the Omicron variant^15^.

### Stratification of vaccine-elicited time-course antibody dynamics

We developed a mathematical model describing the vaccine-elicited antibody dynamics to evaluate the impact of primary two-dose COVID-19 vaccination on rapid immunity at the individual level and reconstructed the best-fit antibody titer curves of 2,407 participants in the Fukushima cohort (details are provided in the **Methods**). We then performed an unsupervised clustering analysis using a random forest dissimilarity to stratify the time-course patterns of antibody dynamics into six groups (i.e., G1 to G6) (details in **Supplementary Note 2**). **Fig 2A** represents a two-dimensional Uniform Manifold Approximation and Projection (UMAP) embedding of these six groups, which clearly shows that G5 and G6 are separated from the other groups. Using a different color for each group, we also plotted the reconstructed individual antibody dynamics after 25 days after the first vaccination as shown in **Fig 2B** (see also **Extended Data Fig 2D**). The projected time course of each group showed that sustained antibody titers remained high among individuals in G1 (i.e., the “rich” responder group), whereas those in G5 and G6 were low (i.e., “poor” responders) (**Fig 2C**). We classified G1 as a “durable” population and G5/G6 as a “vulnerable” population in our analysis. In contrast, individuals in G2, G3, and G4 showed intermediate dynamics. Note that most individuals in the Fukushima cohort had no history of natural infection (i.e., only 1.07%).

**Figure 2.**
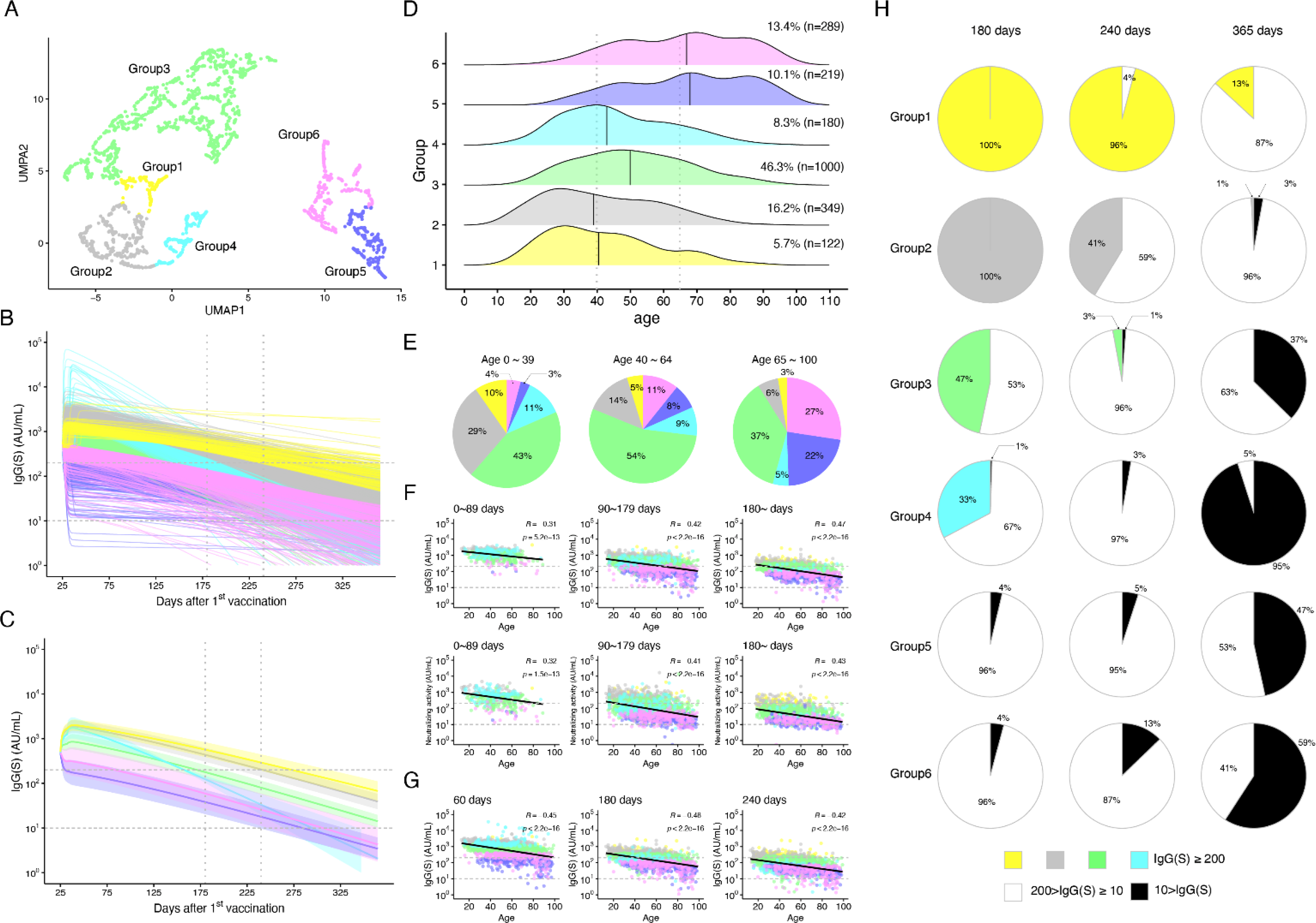
Stratification of vaccine-elicited antibody response. **(A)** UMAP of stratified antibody responses to mRNA vaccination based on the extracted features from the reconstructed individual-level antibody dynamics are shown. Data points represent individual participants and are colored according to group (i.e., G1 to G6). **(B)** The reconstructed individual antibody dynamics after 25 days after the first vaccination in each group are shown. **(C)** The time-course patterns highlighted by Partial Least-Squares Discriminant Analysis (PLS-DA), which discriminates each group from the others, are shown. The individual dynamics after 25 days after the first vaccination were projected onto the direction of the first latent variable. The horizontal dashed lines at 200 and 10 AU/mL correspond to the thresholds of “vaccine efficacy” and unvaccinated persons, respectively. The vertical dotted lines represent 180 and 240 days after the first vaccination. **(D)** Age distribution in each group along with its fraction (%) and the number of individuals (n) is shown. **(E)** Group distribution by age (i.e., 0-39, 40-64, 65-100) is plotted in pie charts. **(F)** Measured IgG(S) and neutralizing activity in each group are plotted in the early, middle, and late period. **(G)** Simulated IgG(S) titers from reconstructed individual-level antibody dynamics at 60, 180, and 240 days after the first vaccinations are plotted. **(H)** Fraction of individuals with antibody titers <10, 10-200, and 200 < AU/mL in each group at 180, 240, and 365 days are plotted as black, white, and group color, respectively, in pie charts. All correlations were calculated as Pearson correlation coefficients.

Next, we investigated the age dependency of the 6 groups (**Fig 2D**): G1/G2, G3/G4, and G5/G6 consisted mainly of people aged 0 to 39 years (young), 40 to 64 years (middle age) and 65 to 100 years (old), respectively. Most individuals were stratified into G3 or G2, the representative population of the Fukushima cohort (**Fig 2E**). Although the age distributions of G1/G2 and G3/G4 were similar, individuals in G2 and G4 showed a relatively rapid decay in antibody response after the second dose compared with those in G1 and G3, respectively (**Extended Data Fig 2D**).

The longitudinal IgG(S) and neutralizing activity measured by CLIA are separately plotted into three different periods (i.e., 0-89, 90-179, ≥180 days) by taking into account the stratified groups in **Fig 2F**. Both IgG(S) and neutralizing activity decreased as age increased and time passed. More precise predictions (i.e., at a specific date at an individual level rather than during a specific period at a population level) were made possible with our reconstructed individual-level antibody dynamics (**Fig 2G**).

To further evaluate the vaccine-elicited immunity, we calculated the fraction of individuals with antibody titers of IgG(S) below set thresholds (i.e., 10 and 200 arbitrary unit/mL [AU/mL]) (**Fig 2H**). As reported in the study by Nakano et al.^25^, 10 AU/mL is a clear threshold for unvaccinated individuals, meaning that individuals showing titers less than this threshold have no vaccine efficacy at all. In contrast, so far, there are no clear thresholds for quantifying vaccine efficacy, as intensively discussed in Cromer et al.^2^ and Khoury et al.^3^. Here we used the value of 200 AU/mL as an indicator for the efficacy threshold, because more than 80% of vaccinated persons maintained their antibody titers above 200 AU/mL during the early period (i.e., for at most 3 months after the first vaccination) regardless of group in our cohort (see the top-left panel in **Fig 2F**). Our simulations showed that most of the durable population (i.e., G1) maintained their vaccine efficacy for at least 8 months, and furthermore, around 13% of them still did not show a loss of vaccine efficacy for 1 year after the first vaccination (**Fig 2H**). On the other hand, the vulnerable populations (i.e., G5/G6) showed a low induction of antibody titers after the second dose and rapid decay in efficacy; approximately 50% of them had very low antibody titers after 1 year, which were comparable to those in unvaccinated persons. Individuals in G4 showed a high initial induction of antibody titers after the second dose but a rapid decay; approximately 95% of them showed a loss of vaccine efficacy after 1 year.

For our sensitivity analysis, we used the threshold of 80 AU/mL, which corresponds to “strong positive” as defined in the Mount Sinai Health System^26^ and obtained the same conclusions (e.g., **Supplementary Fig 1A** for 80 AU/mL).

### Characterizing vulnerable and durable populations

Multiple factors, including underlying medical conditions, drug history, and immune competency, may contribute to the stratification of the study population. For example, there was substantial variation in sustained immunity within the same age group (e.g., age distributions in both the durable and vulnerable populations were wide), and about 7% of the vulnerable population in G5/G6 were young individuals (0-39 years of age) (**Fig 2E**).

To further characterize each stratified group, we trained a random forest classifier to predict the group from the sample characteristics of 2159 individuals in the Fukushima cohort (see **Extended Data Table 1**), and obtained ROC-AUCs of 64.4%, 69.3%, and 70.9% for predicting G1 (the durable population), G2, and G1/G2, respectively (**Fig 3A**). Analysis of the feature importance suggested that adverse reactions (e.g., headache and joint pain) were significant predictors common to both G1 and G2 (**Fig 3C**). On the other hand, we achieved ROC-AUCs of 78.0%, 75.1%, and 80.8% for predicting G5, G6, and G5/G6 (the vulnerable populations), respectively (**Fig 3B**). Various factors including age, the interval between the two doses, comorbidities, and medication use were listed as the significant predictors of G5 and/or G6 (**Fig 3D**). This finding was consistent with the results of our literature review (see **Supplementary Table 3**). We were not able to achieve a high ROC-AUC for predicting G3 and G4, the groups into which the majority of our cohort participants fell (see **Extended Data Fig 3A** and **Discussion**). Furthermore, when the classifier for G5/G6 was trained after the population was stratified according to age (i.e., 0-39 [young], 40-64 [middle age], and 65-100 [old]), the ROC-AUCs were 69.3%, 68.7%, and 70.1%, respectively (see **Extended Data Fig 3B**). Our analysis of the feature importance suggested that medication use (e.g., anti-cancer agents, steroids, and immunosuppressants) was a significant predictor for young people, whereas alcohol consumption and smoking played an additional role for middle-aged persons (**Fig 3E**). The detailed inter-relationship of the various features described in **Fig 3F** and **Extended Data Fig 3C** are explained in **Supplementary Note 3**.

**Figure 3.**
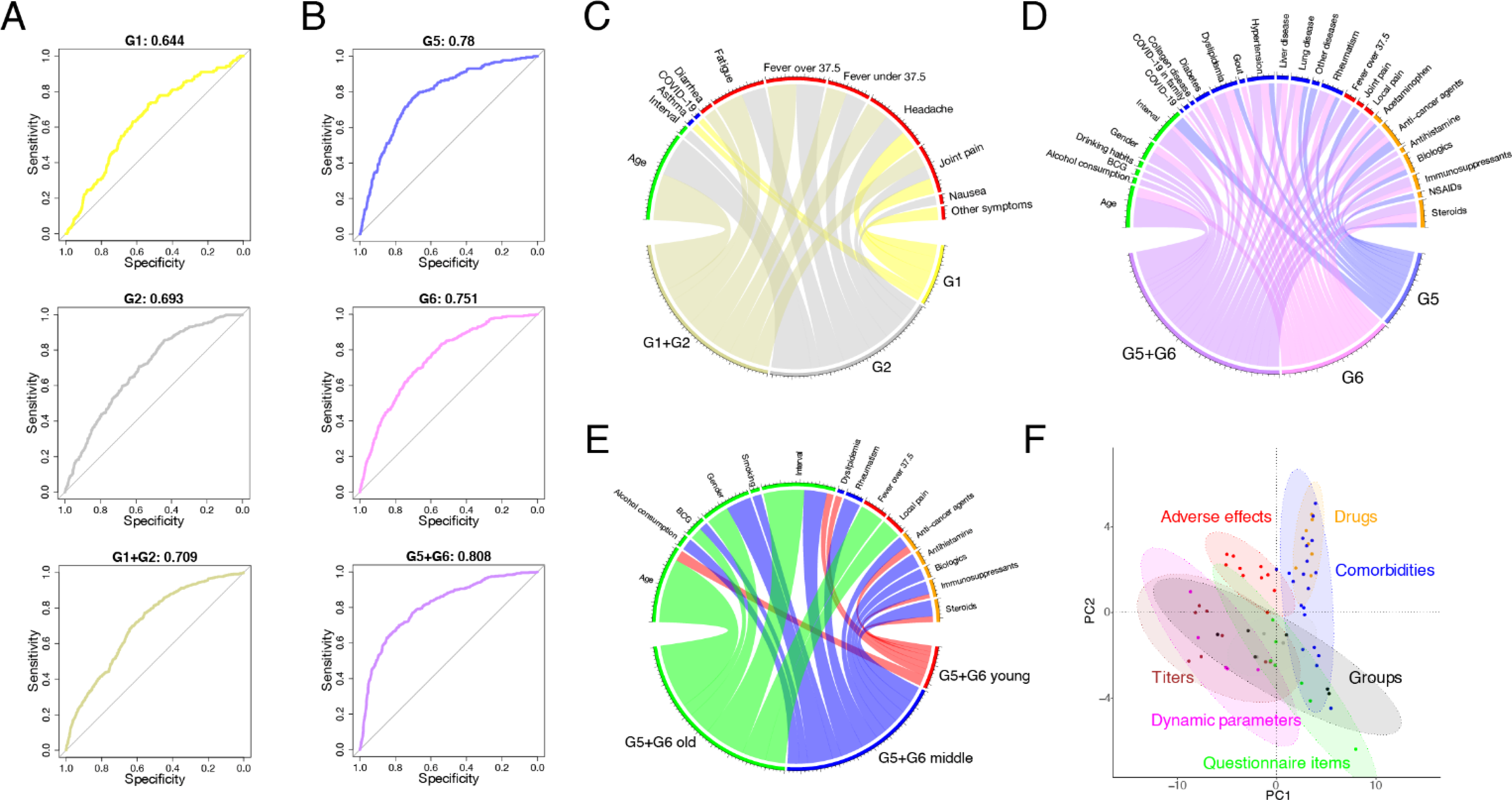
Characterization of vulnerable and durable populations. **(A)** and **(B)** show the ROC curves of random forest classifiers trained on predicting either G1 (durable population); G2 or G1/G2; and G5, G6, or G5/G6 (vulnerable population), respectively. The corresponding AUC value of each ROC curve is calculated on the top of each panel. **(C)** and **(D)** show the most predictive features (feature importance) of G1, G2, and G1/G2 and G5, G6, and G5/G6 as Chord diagrams, respectively. Each feature is represented by a fragment on the outer part of the circular layout. The size of the arc from a group to a feature is proportional to its importance as measured by mean decrease in accuracy. Features with *p* < 0.05 are displayed. **(E)** The most predictive features of G5/G6 in young, middle-aged, and old populations are calculated and shown as a Chord diagram. Features with *p* < 0.05 are displayed. **(F)** Principal component analysis of various features is shown. The colored ellipses represent 90% data ellipses drawn for each category.

### Predicted effect of booster vaccination on stratified antibody dynamics

Several studies have reported that booster doses after the primary two-dose vaccination rapidly increase antibody titers to a high level, and that these booster-elicited antibody responses induce substantial neutralization against VOCs, including the Omicron variant^15–17^. However, it remains unclear how booster vaccinations shape the induction of antibody response depending on the second-dose, vaccine-elicited antibody dynamics (i.e., the time-dependent vaccine-elicited antibody history). Despite the current limited vaccine supply and large variation even within the same stratified group, the priority for booster vaccines at the “individual level” of immunity and the effect of boosters on “population-level” immunity are not well understood.

Assuming similar immune kinetics of the second-dose, vaccine-elicited antibody dynamics, we modeled the elicited antibody responses on each group over the first year after the booster vaccination at 8 months after the first vaccination (**Fig 4A**). Consistent with the current booster vaccination studies^13, 14, 16–20, 27^, our simulations showed a rapid and high induction of antibody responses regardless of group. Moreover, our simulation supported that a third-dose vaccination booster protects against VOCs in a wide range of populations. In contrast, 35% and 9% of G5 and G6, respectively, at 1 month after booster vaccination showed limited induction (i.e., less than 200 AU/mL) (see **Fig 4B**). We also confirmed a similar trend for booster vaccination at 1 year (**Extended Data Fig 4A**), and at the threshold of 80 AU/mL at 8 months (**Supplementary Fig 1B**). The existence of these vulnerable populations may explain the considerable variability in booster-elicited antibody responses observed in several cohort studies^6–8, 15^. Targeting individuals in G5/G6 for booster vaccination is expected to provide protection from breakthrough infection.

**Figure 4.**
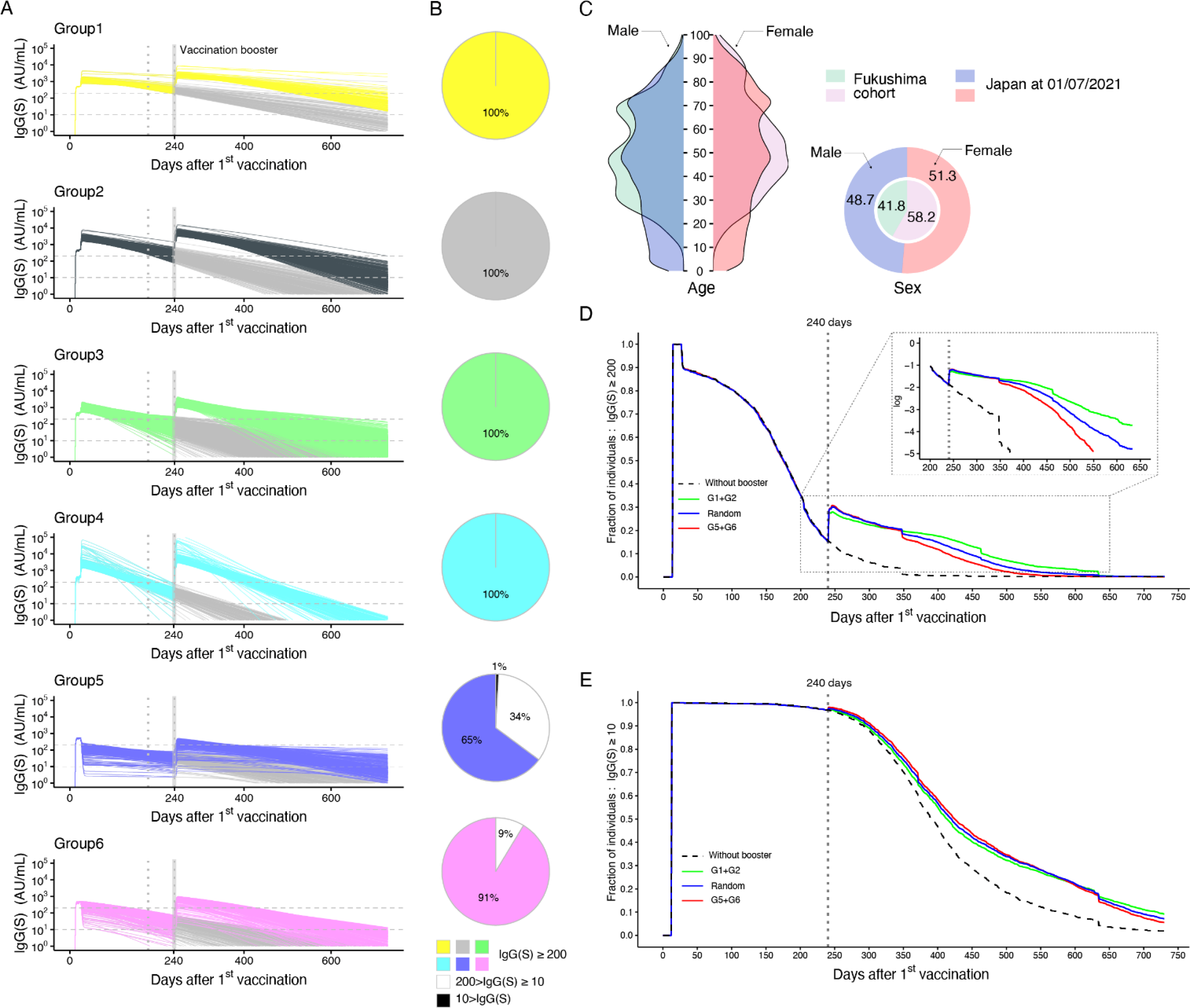
Simulating booster vaccines. **(A)** Predicted individual elicited antibody responses for each group over the first year after booster vaccination at 8 months after the first vaccination are shown. The horizontal dashed and vertical dotted lines correspond to 200 and 10 AU/mL and at 180 and 240 days after the first vaccination, respectively. **(B)** Fraction of individuals with antibody titers of <10, 10-200, and 200 < AU/mL in each group at 1 month after booster vaccination are plotted as black, white, and group color, respectively, in pie charts. **(C)** Age and sex distributions of a hypothetical cohort adjusted via population estimates in Japan are shown along with those in the Fukushima vaccination cohort. **(D)** and **(E)** show the time-dependent fractions of individuals with antibody titers above 200 and 10 AU/mL with (i.e., colored curves) and without (i.e., black dashed curve) 20% booster vaccination at 240 days after the first vaccination, respectively. The green, blue, and red curves correspond to the booster vaccination targeting individuals randomly sampled from G1/G2, G1-G6, and G5/G6, respectively. Enlarged view of the dotted box in **(D)** shows the fraction plotted in log-scale.

Next, taking advantage of our stratification, we further evaluated the effect of booster vaccination on population-level immunity. Here we assumed a hypothetical cohort of 10,000 individuals, adjusted for age and sex distributions according to population estimates in Japan as of 1 July 2021 by the Statistics Bureau of Japan, Ministry of Internal Affairs and Communications^28^ (**Fig 4C**). We excluded populations aged less than 12 years in our simulations to reflect the age threshold of the cohort.

First, we simulated the fraction of individuals with antibody titers below the thresholds (i.e., 10 and 200 AU/mL) when all individuals received a booster vaccine (i.e., 100% booster coverage) at different time periods (8 and 12 months after the first vaccination) in **Extended Data Fig 4BC**. As expected, 100% booster coverage effectively increases and recovers their immune responses regardless of timing compared with the no booster scenario. Since 100% booster coverage is too optimistic given the limited vaccine supply, we next applied more realistic booster strategies: 20% and 50% coverage of booster vaccinations (**Fig 4DE** and **Extended Data Fig 4DE**). We compared the booster vaccinations to 2,000 individuals (i.e., 20% of the total 10,000 population: 20% booster vaccination) in G1/G2 (green curve: the durable population and others), G5/G6 (red curve: the vulnerable population), and randomly sampled groups (blue curve) in **Fig 4DE**. Although small differences were observed for the fraction with the threshold of 200 AU/mL (**Fig 4D**), the booster vaccinations provided similar benefit to population-level immunity regardless of population clusters. Similar trends in 50% booster coverage are shown in **Extended Data Fig 4DE**. Here we again stress that the same conclusions are obtained with different thresholds instead of 200 AU/mL (e.g., **Supplementary Fig 1CDE** for 80 AU/mL). Considering the relative benefit of booster vaccinations at the individual level, our identified vulnerable population is a candidate for priority booster vaccination under the limited vaccine supply (see **Discussion**).

## Discussion

As of May 6, 2022, there were 266 COVID-19 vaccine candidates undergoing phase 1/2 clinical trials and 124 candidates in phase 3 clinical trials (ClinicalTrials.gov). However, it is expected to take more time until second-generation vaccines that block transmission of Omicron and other novel virus variants with a much higher probability than current ones are available around the world. We still need to use the current first-generation vaccines as vaccine boosters. Therefore, determining the priority for booster vaccines in the vulnerable population is an important public health concern given a limited global supply of vaccines^13–20^.

In this study, we aimed to understand the individual-level variation in vaccine-elicited immune responses at the time of booster vaccinations and the effect of booster vaccination. Only 1.07% of our cohort of individuals had a history of natural infection; thus, the influence of natural infection was minimal in the present study. We repeatedly assessed and monitored antibody titers following two doses of a SARS-CoV-2 vaccine in the unique Fukushima cohort of 2,526 individuals.

Previous studies generally investigated differences in vaccine-elicited antibody responses after stratification of the population according to variables including age, sex, lifestyle habits, comorbidities, adverse reactions, and medication use (i.e., in “given” groups). For example, Levin and colleagues explored differences in antibody responses according to age, sex, and immunesupression^8^. Ward and colleagues also showed that antibody responses are significantly decreased in individuals with diabetes, obesity, smoking habits, cancer, immunosuppression, or neurological conditions after adjustment for age and sex^29^. It is well known that active cytotoxic chemotherapy and specific comorbidities such as kidney disease, hypertension, and heart disease significantly affect antibody responses^22, 30–32^. Importantly, in our cohort, we identified six groups with different antibody responses by using a combined approach with process-based mathematical modeling and data-driven analysis (i.e., without determining these “given” groups in advance as mentioned above). In particular, we found that G1 mainly consisted of participants aged 0 to 39 years who were part of the durable population (i.e., those who were “rich” responders showing high sustained antibody titers), whereas G5/G6 consisted of participants aged 65 to 100 years who were part of the vulnerable population (that is, those who were “poor” responders showing low sustained antibody titers). Our study showed that more than 10% of the durable population maintained their IgG(S) titers above 200 AU/mL for 1 year after the first vaccination, whereas approximately 50% of the vulnerable population could not maintain their antibody titers above 10 AU/mL after 1 year (i.e., they completely lost their vaccine efficacy).

An important question is how we maintain both individual- and population-level immunity through booster vaccinations. Although T and B cell responses may contribute to protection from symptomatic and severe infections^4, 12, 27, 33^, recent studies have shown that vaccine efficacy is strongly linked to the elicited antibody responses, meaning that sustained high antibody titers above a certain threshold are crucial^2, 3, 34^. The present study showed that vulnerable populations (i.e., G5/G6) include individuals who are not necessarily older adults nor with high-risk conditions. In fact 5.5% of G5/G6 were young adults who did not have any underlying medical conditions but reported at least one adverse reaction. Although there is no clear consensus for the threshold so far, we found that individuals in G4 maintained high antibody titers induction for months, but their titers rapidly decreased below 200 and 80 AU/mL around 6 and 7 months after the first dose on average, respectively. Because individuals who are classified in G4, G5, and G6 represent those with poor or rapidly decaying vaccine-elicited antibody responses who would not necessarily benefit from the current prioritized booster program on the basis of age and prior medical conditions, it may be important to better understand the large variation in vaccine-induced immunity at the individual level as shown in the present study. Identifying these target individuals and recruiting them to booster vaccinations are important tactics.

Most of the durable population (i.e., G1) will maintain their antibody titers induced by the second dose of mRNA vaccine at high levels at least for 8 months after the first dose, implying that the priority for booster vaccinations is low. Individuals in G1 had no history of natural infection except for one participant. In our study cohort, G1 made up only 5.7% of the cohort, and we only identified these participants because of the large participant size of the Fukushima cohort (over 2,500 individuals). In other words, if the cohort size was in the range of several hundreds, which is the usual size of prior studies made up of health care workers volunteers^30, 35–38^ (e.g., 93.5% of 93 cohort studies in our literature review had less than 1,000 participants, see **Supplementary Note 1**), these durable individuals might not have been identified and might have been overlooked as outliers (e.g., around 5 individuals in a cohort of 100 individuals). Although the representativeness of the small cohort is unclear (only 61 individuals), preexisting high antibody titers before booster vaccination may limit the extent of antibody boosting (see below)^15^.

Our study suggests that booster vaccination of the vulnerable populations of G5/G6 at 80 days after the first vaccination increases antibody titers at both the individual and population level. Booster vaccinations added similar effects to population-level immunity regardless of a population profile under the limited vaccine supply (i.e., 20% and 50% booster coverage) in our simulations. These scenario-based simulations provide novel insights on the timing for booster vaccinations to maintain both individual- and population-level immunity against the emerging and future VOCs.

Overall, our analysis showed a large variation in vaccine-elicited antibody response at the individual level^6–8^. Additional work is required to evaluate how specific antibody neutralizations of VOCs, as quantified by live virus, influence the stratification^13, 15, 27^, and how vaccine-elicited cellular immunity, including B and T cell responses, would explain such variations^12, 15^. In particular, we used only the basic demographic information for the characterization of the six groups and are currently collecting additional longitudinal immunologic information, such as cellular immune changes (e.g., memory B cell compartments) and cytokine and chemokine profiles of the vaccinated individuals in our cohort. This will enable us to test and further characterize and understand durability and vulnerability in G1 and G5/G6, respectively^15^.

There is an assumption in our mathematical model underlying the vaccine-elicited antibody response: antibodies are simply assumed to be generated from one compartment of B cells including heterogeneous cell populations that produce antibodies. That is, our mathematical model describes the vaccine-elicited antibody responses induced by the average B cell populations. As additional datasets become available on time course and quantitative phenotypic B cell populations such as short-lived antibody-secreting cells (i.e., plasmablasts), long-lived antibody-secreting cells (i.e., plasma cells), and SARS-CoV-2-specific (class-switched) memory B cells,^9, 39^ it will be necessary to adjust our mathematical model to more precisely differentiate the antibody dynamics of different B cells. In a recent study with a small sample size, the decay rate of S-specific IgG from its peak to around 6 months after the first dose was similar, but the average decay slowed between 6 and 9 months^15^. Other studies suggest that germinal center B cell responses are critical to maintain the longevity of sustained antibody titers apart from the quick and large antibody production immediately after the second vaccination (or infection)^40–42^. Although this emerging evidence for the heterogeneity of B cell populations implies that the two-compartment model of antibody-secreting cells (i.e., plasmablasts and plasma cells) may better describe IgG(S) dynamics, our mathematical model describes the IgG(S) dynamics of 2,159 individuals (of which 208 individuals had 3 samples collected) at least until around 9 months after the first dose. Our stratification is based on feature engineering but not reconstructed antibody dynamics directly, meaning that the effect of the late-phase dynamics is minimal. However, long-term prediction of antibody dynamics by use of our mathematical model will not be possible.

Important research questions remain: whether people will be classified into the same (or similar) stratified groups after booster vaccination, how the booster vaccine-elicited antibody response will change depending on the interval between the second and third doses, and which group shows low and high neutralization to (emerging) VOCs, for example. The Fukushima cohort is an ideal environment in which to evaluate how immunity, including humoral and cellular responses of durable and vulnerable individuals, is increased by future SARS-CoV-2 infections after the booster vaccination. Taking advantage of our Fukushima cohort, we will further evaluate the impact of booster vaccinations and post-vaccination infections on the identified vulnerable populations^43, 44^.

## METHODS

### Ethics statement

The study was approved by the ethics committees of Hirata Central Hospital (number 2021-0611-1) and Fukushima Medical University School of Medicine (number 2021-116). Written informed consent was obtained from all participants individually before the survey.

### Participant recruitment and sample collection

The candidates were mainly recruited from Hirata village, Soma city, and Minamisoma city in rural Fukushima prefecture. We conducted sequential blood sampling and non-sequential blood sampling. Twelve health care workers participated in the sequential blood sampling, and their vaccination and blood sampling schedule are shown in **Fig 1E**. A total of 2526 individuals participated in non-sequential blood sampling. Health care workers, frontline workers, and administrative officers from each municipality were intentionally recruited to keep the cohort size large and the dropout rate low. Although most of the health care workers, frontline workers, and administrative officers were under the age of 65, relatively healthy community-dwelling older adults living in the community and in long-term care facilities were also recruited to maintain a wide age range for the cohort. Blood sampling was performed once during each period in June, September, and November 2021, respectively. The first vaccine dose was administered between March 10 and August 20, 2021, and the second dose between March 31 and September 14. The median (interquartile range) interval for the two-dose vaccination was 21 days. A total of 226 health care workers participated in the first blood sampling between May 31 and June 6, 2021. A total of 2526 individuals participated in the second blood sampling between September 8 and October 8, 2021. A total of 2443 individuals participated in third blood sampling between November 21 and December 25, 2021 (**Fig 1E**). Note that the 12 health care workers (described in **Extended Data Fig 5B**) with sequential blood sampling were not included in the non-sequential population of 2526 participants. In conclusion, of the total 2526 participants, those eligible for analysis were those who completed the second vaccination and at least two blood samplings. Participants whose antibody titer was higher than in the previous blood sampling in non-sequential blood sampling were excluded from the final analysis (**Fig 1B**).

Information on sex, age, daily medication, medical history, date of vaccination, adverse reaction after vaccination, type of vaccination, blood type, bacillus Calmette–Guérin (BCG) vaccine history, smoking habit, and drinking habit was retrieved from the paper-based questionnaire (summarized in **Extended Data Table 1**). All blood sampling was performed at the medical facilities with 8 mL, and serum samples were sent to The University of Tokyo.

### SARS-CoV-2-specific antibody measurement

All serological assays were conducted at The University of Tokyo. Specific IgG (i.e., IgG(S)) and neutralizing activity were measured as the humoral immune status after COVID-19 vaccination. Specific IgG antibody titers (IgG(N)) were used to determine past COVID-19 infection status. Chemiluminescent immunoassay with iFlash 3000 (YHLO Biotech, Shenzhen, China) and iFlash-2019-nCoV series (YHLO Biotech, Shenzhen, China) reagents were used in the present study. The threshold value was 10 AU/mL. The measurement range was 2-3500 AU/mL for IgG(S) and 4-800 AU/mL for neutralizing activity. For neutralizing activity, AU/mL×2.4 was used to convert to International Units (IU/mL); for IgG(S), AU/mL×1.0 was used to convert to binding antibody units (BAU/mL). The testing process was as per the official guideline. Quality checks were conducted every day before starting the measurement.

### Mathematical modeling of (booster) vaccine-elicited antibody dynamics

We developed a simple but quantitative mathematical model describing the vaccine-elicited antibody dynamics as follows:

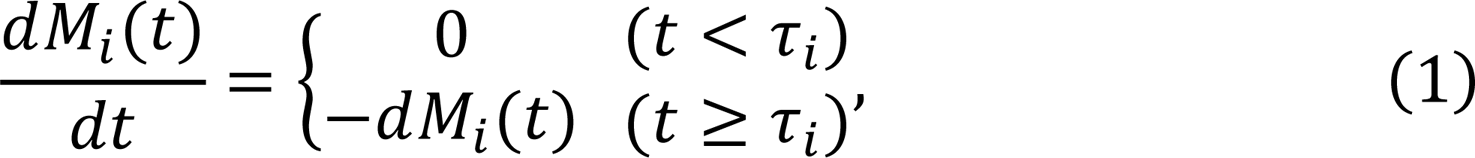

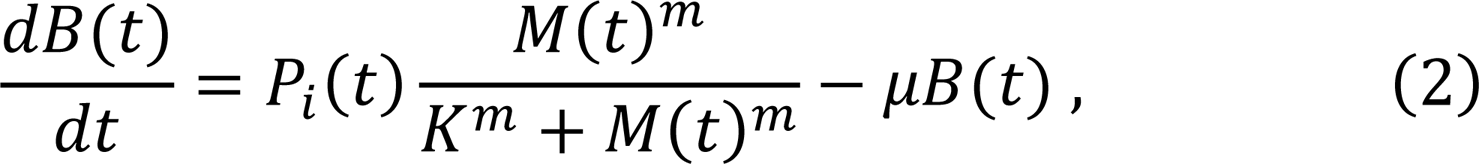

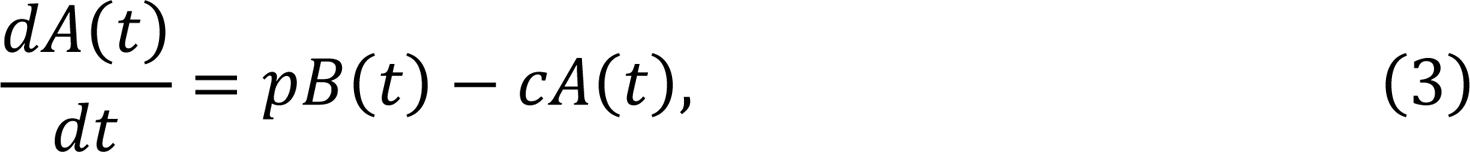

where the variables *M*_*i*_(*t*), *B*(*t*), and *A*(*t*) are the amount of mRNA inoculated by the *i* -th vaccination, the number of antibody-secreting cells, and the antibody titers at time *t*, respectively. Parameters τ_*i*_ and *d* represent the timing of the *i*-th vaccination and the decay rate of mRNA. Note that τ_1_ = 0 is assumed for all individuals and τ_2_ − τ_1_ corresponds to the interval of the first and second vaccinations. We here considered that *D*_*i*_ is the inoculated dose of mRNA by the *i*-th vaccination, that is, *M_i_*(τ_*i*_) = *D*_*i*_ for *i* = 1,2, …, *n*, and 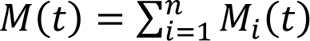.

In our mathematical model, one compartment of B cells including heterogeneous cell populations that produce antibodies is assumed, and therefore the product of *P*_*i*_(*t*) and *M*(*t*)^*m*^/(*M*(*t*)^*m*^ + *K*^*m*^) represents the average *de novo* induction of the antibody-secreting cells, that is, the average B cell population dynamics is modeled. Here *P_i_*(*t*) is a step function defined as *P*_*i*_(*t*) = *P*_*i*_ for τ_*i*_ + *η*_*i*_ ≤ *t* < τ_*i*+1_ + *η_i_*_+1_, where *η*_*i*_ is the delay of induction of the antibody-secreting cells after the *i*-th vaccination: otherwise *P*_*i*_(*t*) = 0. The parameters *m*, *K*, and *μ* correspond to the steepness at which the induction increases with increasing amounts of mRNA (i.e., the hill coefficient), the amount of mRNA satisfying *P*_*i*_/2, and the average decay rate of the antibody-secreting cell compartment, respectively.

For example, after the first vaccination, antigen-specific B cells expand and induce antibody-secreting cells and memory B cells. This rapid immune response regarding expansion and induction is formulated by the function of *P*_1_*M*_1_(*t*)^*m*^/(*M*_1_(*t*)^*m*^ + *K*^*m*^) for *η*_1_ ≤ *t* < τ_2_. On the other hand, after the second vaccination, the memory B cells are reactivated by re-exposure to antigen, and this reactivation induces antibody-secreting cells, increases the number of memory B cells, and further re-establishes B cell memory. The recall B cell responses and their antibody secretion are described by the function of *P*_2_(*M*_1_(*t*) + *M*_2_(*t*))^*m*^/((*M*_1_(*t*) + *M*_2_(*t*))^*m*^ + *K*^*m*^) for τ_2_ + *η*_2_ ≤ *t*. The quantity and quality of memory B cells established by the second vaccination are considered in *P*_2_. In a similar manner, the recall B cell responses by the vaccine booster (i.e., the third dose) are simply described by *P*_3_(*M*_1_(*t*) + *M*_2_(*t*) + *M*_3_(*t*))^*m*^/((*M*_1_(*t*) + *M*_2_(*t*) + *M*_3_(*t*))^*m*^ + *K*^*m*^) for τ_3_ + *η*_3_ ≤ *t*. Note that *P*_2_ = *P*_3_, *η*_2_ = *η*_3_ are assumed in our booster simulations. The other parameters, *p* and *c*, represent the antibody production rate and the clearance rate of antibodies, respectively.

Since the clearance rate of antibody is much larger than the decay of antibody-secreting cells, we made a quasi-steady state assumption, *dA*(*t*)⁄*dt* = 0, and replaced Eq.(3) with *A*(*t*) = *pB*(*t*)⁄*c*. Thus, we obtained the following simplified mathematical model, which we used to analyze the antibody responses in this study (see **Extended Data Fig 5A**):

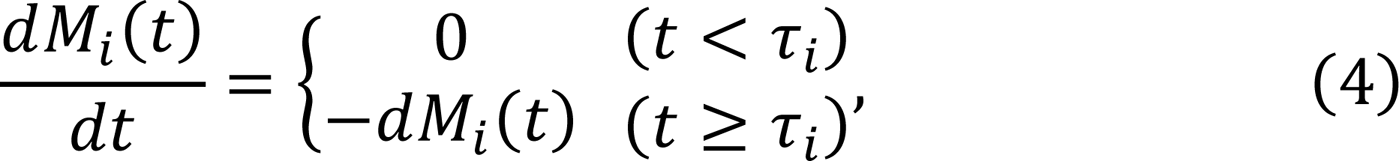

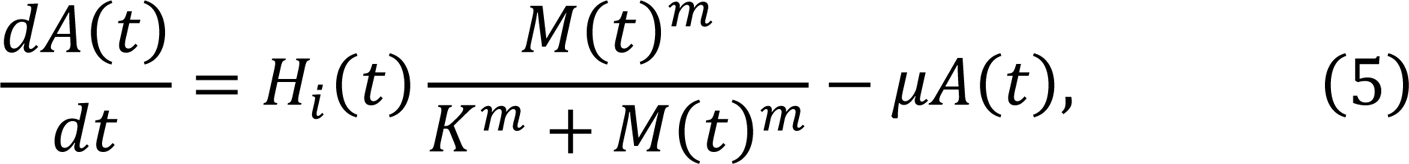

where *H*_*i*_(*t*) = *H*_*i*_ = *pP*_*i*_/*c* for τ_*i*_ + *η*_*i*_ ≤ *t* < τ_*i*+1_ + *η*_*i*+1_. In our analysis, the variable *A*(*t*) corresponds to the IgG(S) titers (AU/mL).

### Quantifying vaccine-elicited time-course antibody dynamics

In addition to the participants in the cohort, we included 12 health care workers whose serum was sequentially sampled for 40 days (on average 25 samples per individual) for validation and parameterization of a mathematical model for vaccine-elicited antibody dynamics. A nonlinear mixed effects model was used to fit the antibody dynamics model, given by Eqs.(4-5), to the longitudinal antibody titers of IgG(S) obtained from the 12 health care workers. The mathematical model included both a fixed effect and a random effect in each parameter. That is, the parameters for individual *k*, *θ_k_*(= *θ* × *e*^*πk*^), are represented as a product of *θ* (a fixed effect) and *e*^*πk*^ (a random effect). *π*_*k*_ follows a normal distribution with mean 0 and standard deviation Ω. We here assumed that the parameters *H*_1_, *H*_2_, *η*_1_, *η*_2_, and *m* varied across individuals, whereas we did not consider interindividual variability in other parameters to ensure parameter identifiability. Note that the half-life of mRNA (i.e., log 2 /*d*) and dose of mRNA (i.e., *D*_*i*_) are assumed to be 1 day^45^ and 100 (*μg*/0.5mL)^46^, respectively. Fixed effect and random effect were estimated by using the stochastic approximation expectation-approximation algorithm and empirical Bayes’ method, respectively. Fitting was performed using MONOLIX 2019R2 (http://www.lixoft.com)^47^. The estimated (fixed and individual) parameters are listed in **Supplementary Table 4**. Interestingly, we found that most of the best-fitted estimated parameters in the mathematical model (i.e., *D*_1_, *D*_2_, *d*, *μ*, *K*, *η*_1_, *η*_2_, *H*_1_) were the same or similar across the 12 individuals compared with those of parameters of *m* and *H*_2_ (see **Supplementary Table 4**). We note that *m* and *H*_2_, which showed wide variation of estimated values, contributed mainly to the vaccine-elicited antibody dynamics after the second vaccination, whereas the other parameters contributed to that after the first dose.

Since 2,407 participants had only 2 or 3 measurements of antibody titers at different time points, we hereafter fixed the parameters in our mathematical model to be the estimated population parameters listed in **Supplementary Table 4**, except *m* and *H*_2_, to accurately reconstruct the large variations in antibody dynamics after the second dose, and these two parameters were independently estimated from each IgG(S) by a nonlinear least-squares method. Although the variation in the 12 individuals may not cover the whole variation of the antibody induction dynamics after the first dose, our analysis focused on those after peak, and the results on the stratification (in particular, the durable and vulnerable populations) are expected to be the same. Using the estimated parameters for each individual, we fully reconstructed the dynamics of IgG(S) titers after the first vaccination (**Extended Data Fig 5B**). We summarized the distribution of parameter values *m* and *H*_2_ for 2,407 participants in **Extended Data Fig 5C**, and the best-fit antibody titer curves of 600 randomly selected individuals are plotted along with the observed data for visualization in **Supplementary Fig 2**.

### Unsupervised clustering and stratification of antibody dynamics

Unsupervised random forest clustering was performed on the selected features of the vaccine-elicited time-course antibody dynamics (rfUtilities package in R). After a random forest dissimilarity (i.e., the distance matrix between all pairs of samples) was obtained, it was visualized with Uniform Manifold Approximation and Projection (UMAP) in a two-dimensional plane and was stratified with spectral clustering (Python scikit-learn). The optimal number of clusters was determined by the eigengap heuristic method.

### Random forest classifiers for characterizing stratified groups

Random forest classifiers were trained to predict either of the six stratified groups (G1-G6) or their combination (G1/G2, G5/G6) using randomForest and rfPermute packages in R. The receiver operating characteristic (ROC) curve for each classifier was drawn from out-of-bag (OOB) samples using the pROC package in R. Feature importance was based on MeanDecreaseAccuracy and was shown as Chord diagrams (circlize package in R). Only the features with *p* < 0.05 (obtained with 1,000 permutations) were selected.

### Statistical analysis

The paper-based questionnaires collected from 2,159 participants were converted into a set of categorical and numerical variables. Numerical variables included age and the interval between the two doses. These variables were then used as input features to random forest classifiers to predict each stratified group (G1-G6). Missing values of categorical variables were treated as a separate category and included in the analyses. The variables used here belonged to any of the following five categories: (i) basic demographic information and lifestyle habits, (ii) information on vaccinations, (iii) underlying medical conditions, (iv) adverse reactions, and (v) medications being taken. As a subanalysis, we also divided participants into three generations according to age (0-39 [young], 40-64 [middle-aged], and 65-100 [old]) and trained random forest classifiers in each generation. When necessary, the same variables were compared among different generations or different groups using Pearson’s chi-square test (for categorical variables), analysis of variance (ANOVA, for more than two numerical variables), or Welch T-test (for two numerical variables). The Pearson correlation coefficient was calculated to evaluate the association between a pair of continuous variables. To calculate the correlation matrix between features, we augmented the variables by adding the following three categories to the above five categories: (vi) antibody titers of each individual, (vii) dynamic parameters of each individual, and (viii) dummy variables specifying each stratified group (G1 to G6). The correlation matrix of these variables (features) was used as an input to principal component analysis. All statistical analyses were performed using R (version 4.1.2).

## Data Availability

All data produced in the present study are available upon reasonable request to the authors

## ACKNOWLEDGMENTS

We would like to thank all the staff from Fukushima Medical University, Seireikai health care group, Hirata village office, Soma City office, Soma Central Hospital, Soma General Hospital, Minamisoma City office, Minamisoma City Medical Association, Minamisoma Municipal General Hospital, Shindo Clinic and Medical Governance Institute, who contributed significantly to the accomplishment of this research, especially, Ms. Yuka Harada, Ms. Serina Noji, Ms. Naomi Ito, Dr. Makoto Kosaka, Mr. Anju Murayama, Mr. Sota Sugiura, Mr. Manato Tanaka, Ms. Yuna Uchi, Mr. Yudai Kaneda, Mr. Masahiko Nihei, Mr. Hideo Sato, Ms. Rie Yanai, Ms. Yasuko Suzuki, Ms. Keiko Abe, Dr. Hidekiyo Tachiya, Mr. Kouki Nakatsuka, Dr. Ryuzaburo Shineha, Ms. Miki Sato, Dr. Masahiko Sato, Mr. Naoharu Tadano, Mr. Kazuo Momma, Mr. Shu-ichi Mori, Ms. Saori Yoshisato, Ms. Katsuko Onoda, Mr. Satoshi Kowata, Mr. Masatsugu Tanaki, Dr. Tomoyoshi Oikawa, Dr. Joji Shindo, Ms. Xujin Zhu, Ms. Asaka Saito, Ms. Yuumi Kondo, and Ms. Tomoyo Nishimura. This study was supported by Medical & Biological Laboratories Co., Ltd., Shenzhen YHLO Biotech Co., Ltd., the distributor and manufacturer of the antibody measurement system (iFlash 3000), Research Center for Advanced Science and Technology in the University of Tokyo, and in part by a National Research Foundation of Korea (NRF) grant funded by the Korea government (MSIT) (2022R1C1C2003637) (to K.S.K.); a Grant-in-Aid for JSPS Scientific Research (KAKENHI) B 18H01139 (to S.I.), 16H04845 (to S.I.), Scientific Research in Innovative Areas 20H05042 (to S.I.); AMED CREST 19gm1310002 (to S.I.); AMED Development of Vaccines for the Novel Coronavirus Disease, 21nf0101638s0201 (to S.I.), 21nf0101638h0001 (to M.T.); AMED Japan Program for Infectious Diseases Research and Infrastructure, 20wm0325007h0001 (to S.I.), 20wm0325004s0201 (to S.I.), 20wm0325012s0301 (to S.I.), 20wm0325015s0301 (to S.I.); AMED Research Program on HIV/AIDS 22fk0410052s0401 (to S.I.); AMED Research Program on Emerging and Re-emerging Infectious Diseases 20fk0108140s0801 (to S.I.); AMED Program for Basic and Clinical Research on Hepatitis 21fk0210094 (to S.I.); AMED Program on the Innovative Development and the Application of New Drugs for Hepatitis B 22fk0310504h0501 (to S.I.); AMED Strategic Research Program for Brain Sciences 22wm0425011s0302; AMED JP22dm0307009 (to K.A.); JST MIRAI JPMJMI22G1 (to S.I.); Moonshot R&D JPMJMS2021 (to K.A. and S.I.) and JPMJMS2025 (to S.I.); Institute of AI and Beyond at the University of Tokyo (to K.A.); Mitsui Life Social Welfare Foundation (to S.I.); Shin-Nihon of Advanced Medical Research (to S.I.); Suzuken Memorial Foundation (to S.I.); Life Science Foundation of Japan (to S.I.); SECOM Science and Technology Foundation (to S.I.); The Japan Prize Foundation (to S.I.); Foundation of Kinoshita Memorial Enterprise (to S.I.); and Kowa Co (to M.T.).

## AUTHOR CONTRIBUTIONS

SI and MT designed the research. YKobashi, YS, TZ, YN, FO, MTa, CY, KS, and MTs conducted the data collection and curation. YT, MK, MY, TA, YS, SN, TK, AS, AN, YS and YKaneko carried out the investigations. NN, KSK, DT, MA, SIwanami, KA, and SI carried out the computational analysis. SI and MT supervised the project. All authors contributed to writing the manuscript.

## INSTITUTIONAL REVIEW BOARD STATEMENT

This study was approved by the ethics committees of Hirata Central Hospital (number 2021-0611-1) and Fukushima Medical University (number 2021-116). This study was conducted in accordance with the Code of Ethics of the World Medical Association (Declaration of Helsinki).

## INFORMED CONSENT STATEMENT

Informed consent was obtained from all subjects involved in the study.

## Supplementary Information

**Supplementary Note 1: Literature review**

This comprehensive literature review aimed to characterize the Fukushima vaccination cohort from previous cohort studies and to identify the factors contributing to the vulnerability of certain populations. A total of 440 articles published up to March 23, 2022, were collected using PubMed. Details of the search terms used in this analysis are provided in **Supplementary Table 1**. In the first step, a group of two or three authors (YT, MK, MY, TA, YS) screened and retrieved articles according to the following two criteria: (i) report evaluating antibody titer testing of human blood samples after two doses of SARS-CoV-2 vaccine; (ii) report collecting at least two blood samples after two doses of SARS-CoV-2 vaccine, and 92 articles met the criteria. Fig 1F shows the overall distribution of the cohort size and study period of the included papers. *Study period* was defined as the maximum period between the first vaccination and the completion of the last sampling, and *cohort size* was defined as the maximum number of participants who had at least one sample collected at certain sampling times during the period. The median cohort size was 103 and the median study period was 150 days, and six articles were found to have a cohort size of 1,000 or more (**Supplementary Table 2**). Next, we extracted and analyzed the factors contributing to vulnerable population status from 40 of 92 papers, including interview sheets, 6 of which were excluded because they did not analyze factors of vulnerability (**Supplementary Table 3**). From this literature review, we found that only the Fukushima cohort (1) consecutively sampled more than 2,000 of the same individuals with less dropout (only 3.3%); (2) included an interview sheet for all participants; (3) targeted “communities” (including non-HCWs); and (4) measured several modalities of antibody titers including neutralizing activity (**Supplementary Table 2**).

**Supplementary Note 2: Stratifying time-course pattern of antibody dynamics**

For the purpose of stratification of the individual vaccine-elicited antibody response, we first applied unsupervised random forest clustering to the individual “reconstructed” antibody dynamics of 2,407 participants (e.g., **Supplementary Fig 2**) but failed to divide the time-course pattern of antibody dynamics into different clusters (data not shown). To overcome this problem, we employed an idea of “feature engineering”: extracting features to improve the quality of results from a machine-learning process, compared with supplying only the raw data to it. We quantified the peak, duration, and area under the curve of the reconstructed antibody dynamics as their features (**Extended Data Fig 5D**). Interestingly, the unsupervised random forest clustering based on these features in addition to our estimated individual parameters (i.e., *m* and *H*_2_) identified 10 clusters that clearly discriminated the time-course patterns (**Extended Data Fig 2AB**). Since the time-course patterns of clusters 2, 3, 6, and 7 were similar and they were close together in two-dimensional Uniform Manifold Approximation and Projection (UMAP) embeddings (**Extended Data Fig 2A**), we merged these clusters into one group. In addition, we removed cluster 8 from further evaluation (i.e., we removed 245 individuals, around 10%, of the 2,407 participants in the Fukushima cohort) on the grounds that their reconstructed antibody dynamics with estimated parameters may not be reasonable. This is because their antibody titers measured from two blood samples showed statistically significantly small differences (*p* < 1.0 × 10^−1^^6^ by Welch two sample t-test) and their sampling intervals were significantly shorter than the others (*p* = 0.01 by Welch two sample t-test) (**Extended Data Fig 2C**). This problem will be solved by adjusting for the timing of their blood sampling in our future study of booster vaccinations. Finally, we obtained 6 groups to evaluate the stratification in further detail (Fig 2A).

**Supplementary Note 3: Inter-relationship of various features**

We visualized the inter-relationship of various features using principal component analysis (PCA) based on their correlation matrix (Fig 3F and **Extended Data Fig 3C**). We found that the first principal axis, explaining 25.9% of the variance, represents titers: the antibody titers and the dynamic parameters of each individual (i.e., their peak, duration, and area under the curve, *m* and *H*_2_) were located in the third quadrant of this PCA plane. The adverse events were in the second quadrant, suggesting their closeness to high antibody titers. However, most questionnaire items about basic demographic information and lifestyle habits remained in the middle, indicating their relatively minor role. A notable exception was age, which was inversely correlated with titers and was indeed situated in the fourth quadrant. By contrast, the second principal axis, explaining 9.8% of the variance, represents comorbidities and medication. Our stratification combines these information and places G1, G2, and G4 in the third quadrant; G3 in the middle; and G5 and G6 in the fourth quadrant near the comorbidities cluster and age.

### EXTENDED DATA FIGURES

**Extended Data Figure 1.**
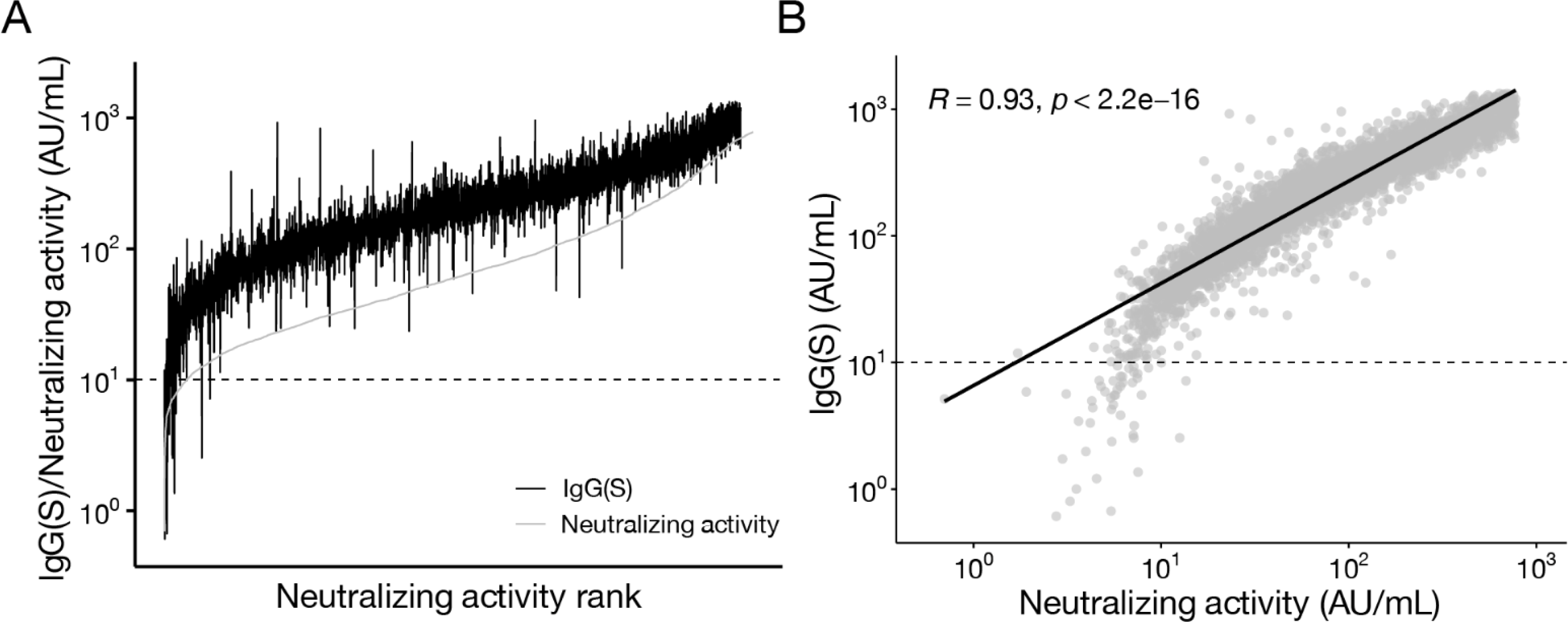
Comparison between IgG(S) and neutralization activity. **(A)** Curve with IgG(S) and neutralization activity on the *Y*-axis and its corresponding neutralization activity rank on the *X*-axis are plotted in black and gray, respectively. **(B)** Correlations between IgG(S) and neutralization activity from same samples are described. Data points represent individual samples. Correlations were calculated as Pearson correlation coefficients.

**Extended Data Figure 2.**
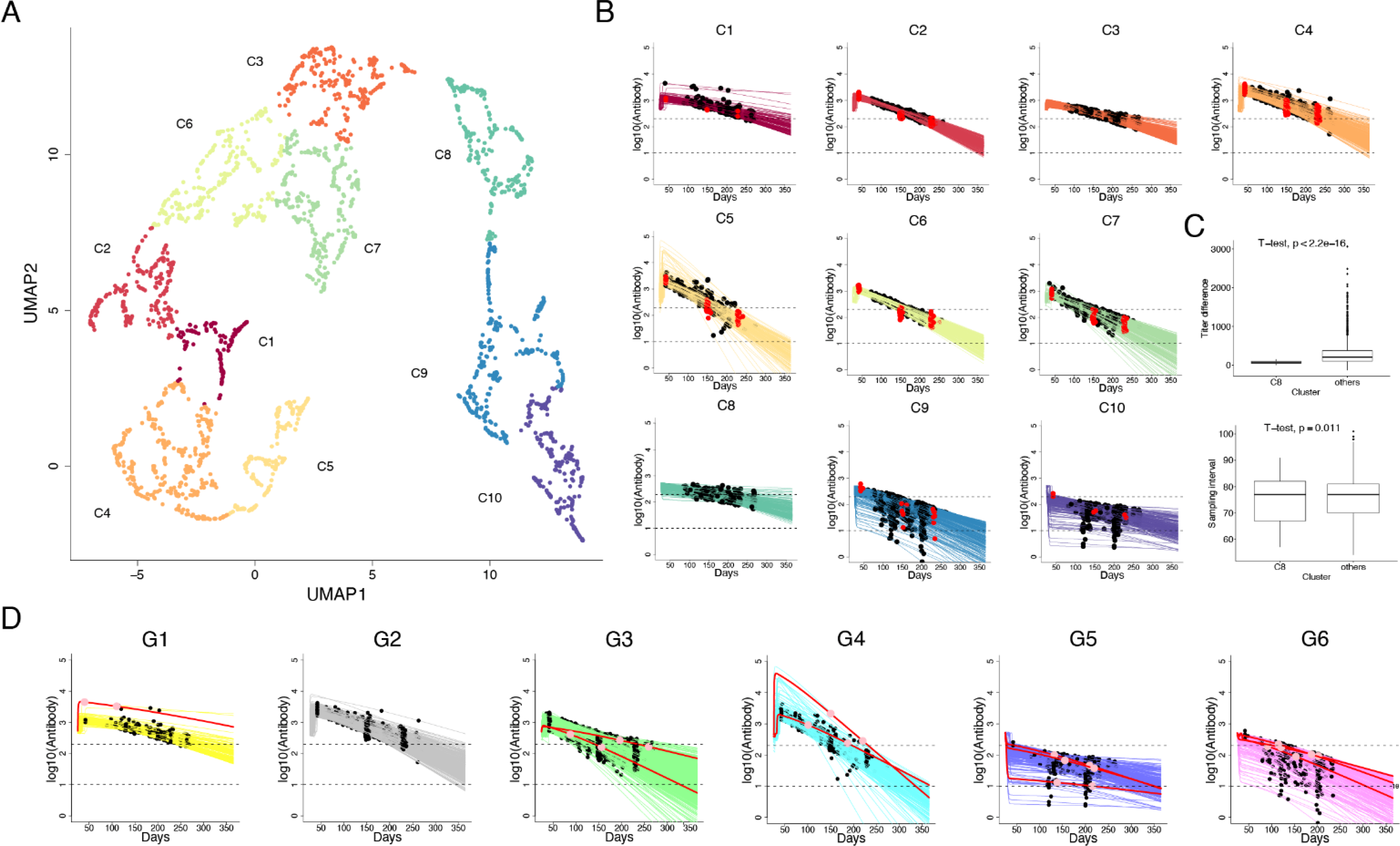
Clustering vaccine-elicited antibody response. **(A)** UMAP of 10 clustered antibody responses based on the extracted features from the reconstructed individual-level antibody dynamics are shown. Data points represent individual participants and are colored by the 10 clusters (i.e., C1 to C10). **(B)** Reconstructed individual antibody dynamics after 25 days after the first vaccination in each cluster in the different colors are represented along with the measured IgG(S). The black and red circles correspond to individuals who had 2 or 3 measurements of antibody titers at different time points, respectively. **(C)** The difference in antibody titers between the two measurements and its sampling interval are compared among clusters. **(D)** Reconstructed individual antibody dynamics after 25 days after the first vaccination in each group in the different colors are represented along with the measured IgG(S) in black and red corresponding to individuals who had 2 or 3 measurements, respectively. The red curves indicate individuals who were infected with SARS-CoV-2 or whose family was infected. The horizontal dashed lines correspond to 200 and 10 AU/mL, respectively.

**Extended Data Figure 3.**
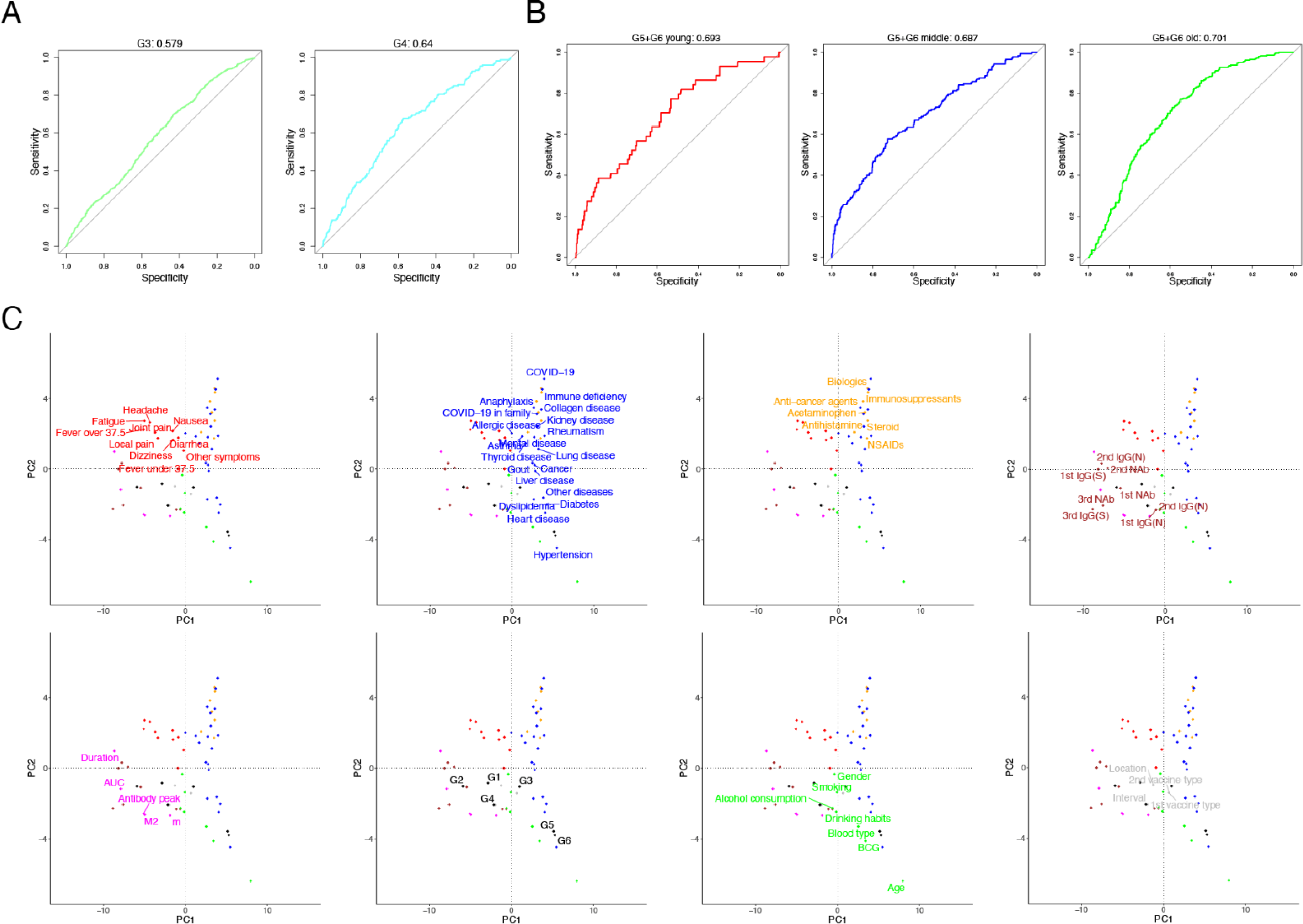
Characterization of stratification. **(A)** and **(B)** The ROC curves of random forest classifiers trained on predicting G3, G4, or G5/G6 in young, middle-aged, and old populations are shown, respectively. The corresponding AUC value of each ROC curve is calculated on the top of each panel. **(C)** Name labels of Figure 3D (principal component analysis of features) are shown.

**Extended Data Figure 4.**
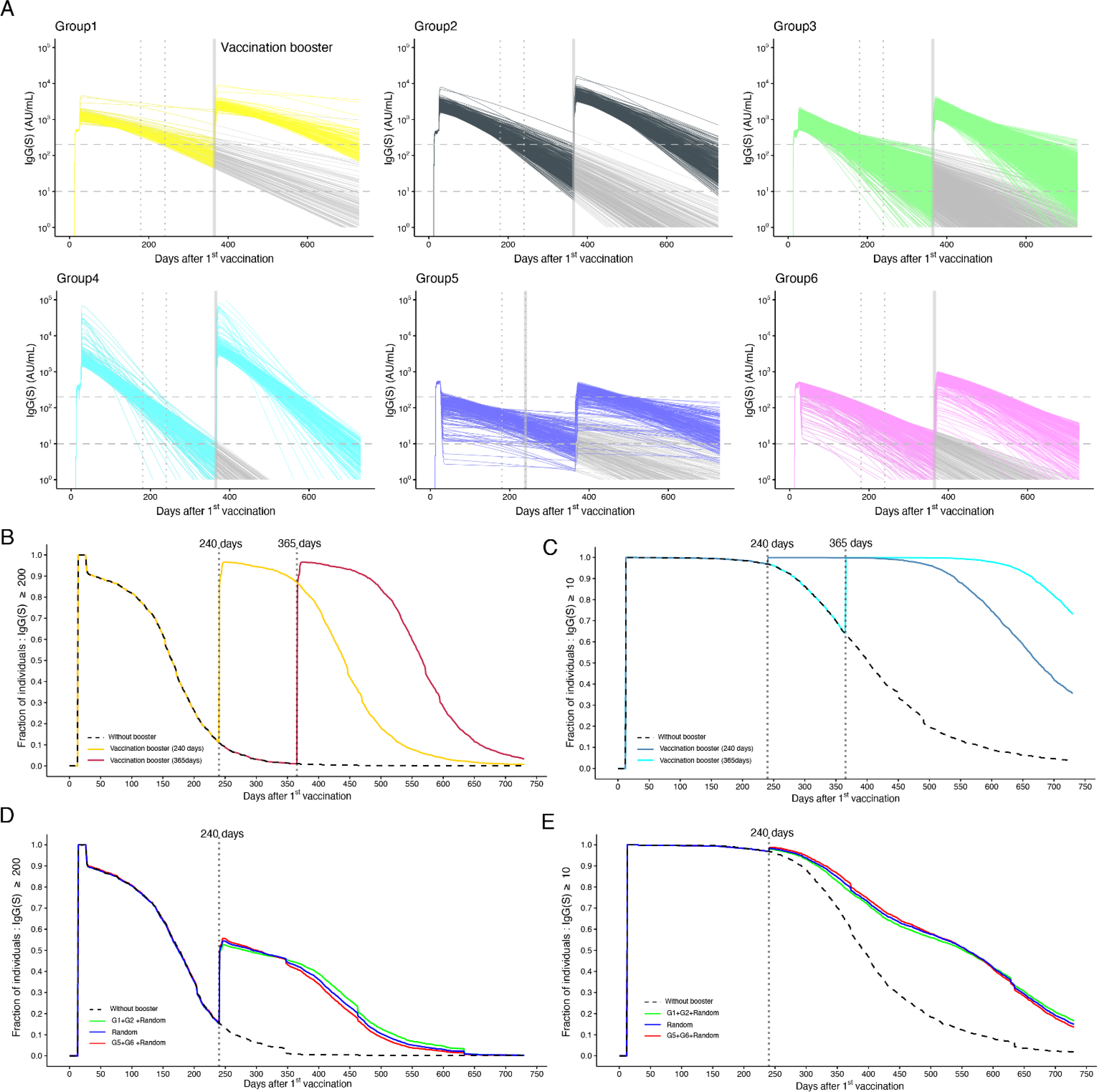
Simulating booster vaccines under different scenarios. **(A)** Predicted individual elicited antibody responses in each group over the first year after the booster vaccination at 1 year after the first vaccination are shown. The horizontal dashed and vertical dotted lines correspond to 200 and 10 AU/mL and at 180 and 240 days after the first vaccination, respectively. **(B)** and **(C)** show the fractions of individuals with antibody titers above 200 and 10 AU/mL with (i.e., colored curves) and without (i.e., black dashed curve) 100% booster vaccination at 240 and 365 days after the first vaccination, respectively. **(D)** and **(E)** show those above 200 and 10 AU/mL with and without 50% booster vaccination at 240 days after the first vaccination, respectively. The green, blue, and red curves correspond to the booster vaccination targeting individuals randomly sampled from “in addition G1/G2, extra are from G1-G6”, “G1-G6”, and “in addition G5/G6, extra are from G1-G6”, respectively.

**Extended Data Figure 5.**
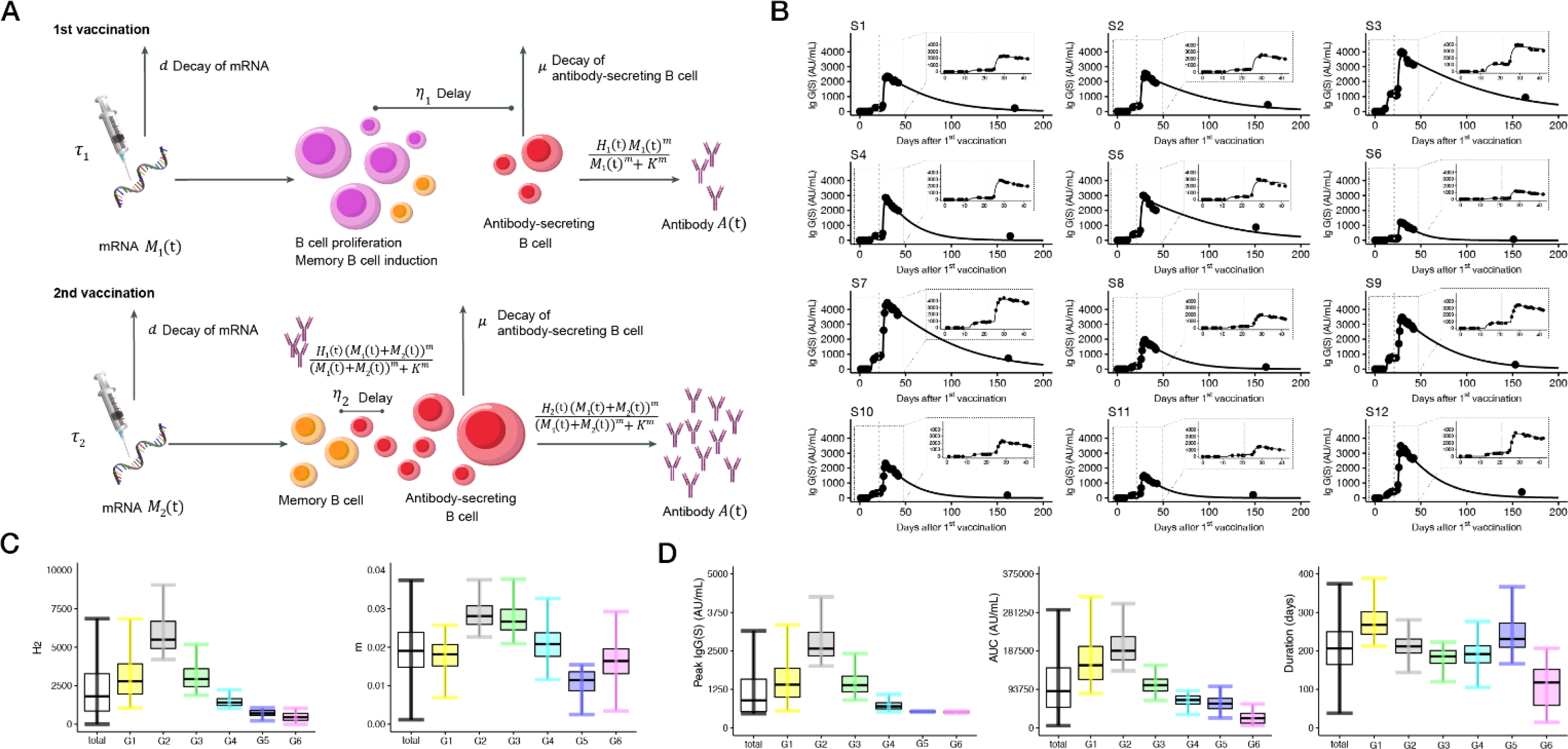
Quantification of vaccine-elicited antibody dynamics. **(A)** Schematic diagram of mathematical modeling. **(B)** Observed and fitted IgG(S) titers for individual participants are described. The IgG(S) titers are obtained from the 12 health care workers with sequentially sampled serum for 40 days (on average 25 samples per individual). Enlarged view of the dotted box shows a fine-level plot focusing on the early period. The dashed vertical lines at day 21 correspond to the date of second vaccination. **(C)** Distributions of estimated parameter values of *H*_2_ and *m* for 2,407 participants are plotted, respectively, among total or stratified populations. **(D)** Distributions of the extracted features from the reconstructed individual-level antibody dynamics are plotted, respectively, among total or stratified populations.

**Supplementary Figure 1.**
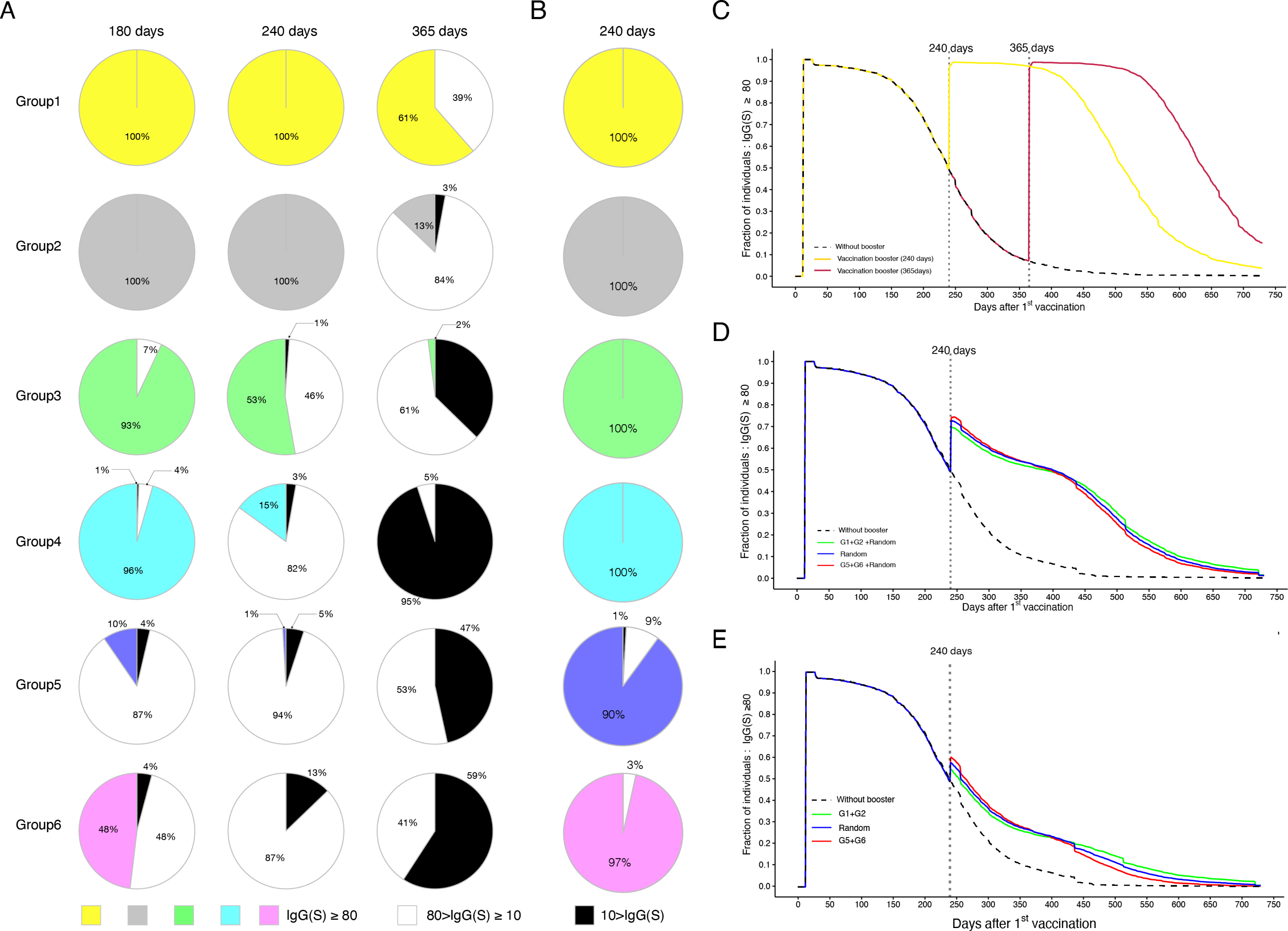
Sensitivity analysis on different thresholds of vaccine efficacy. **(A) and (B)** show the fractions of individuals with antibody titers of <10, 10-80, and 80 < AU/mL in each group at 180, 240, and 365 days after the first vaccination and at 1 month post-booster vaccination plotted as black, white, and group color, respectively (for 200 AU/mL, corresponding to Fig 2H and Fig 4B). **(C)**, **(D)**, and **(E)** represent the time-dependent fractions of individuals with antibody titers above 80 AU/mL with (i.e., colored curves) and without (i.e., black dashed curve) 100%, 50%, and 20% booster vaccination at 240 days (and 365 days for 100% booster vaccination) after the first vaccination, respectively, similar to **Extended Data Fig 4B, Fig 4D** and **Extended Data Fig 4D** for 200 AU/mL of the threshold.

**Supplementary Figure 2.**
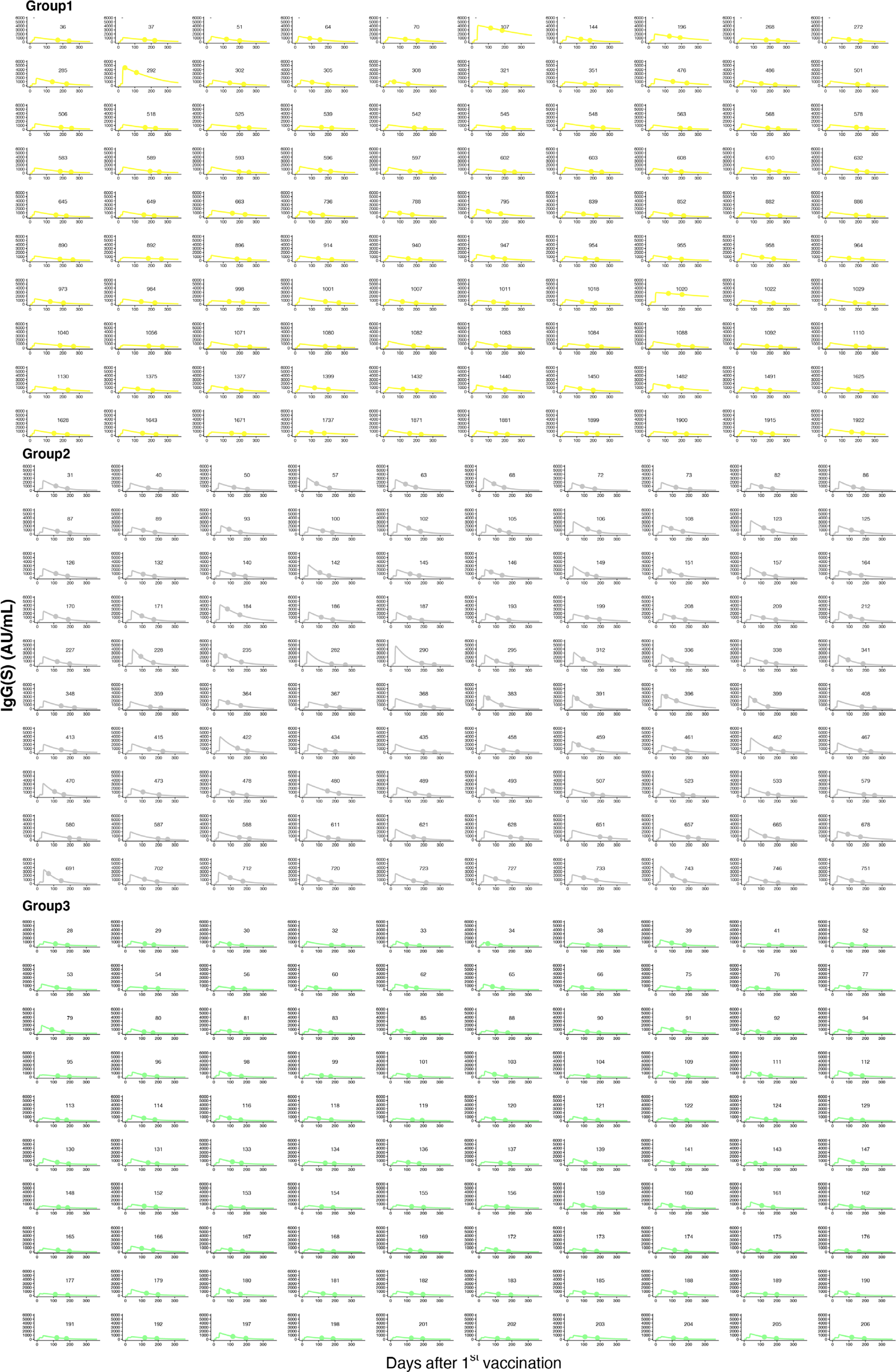
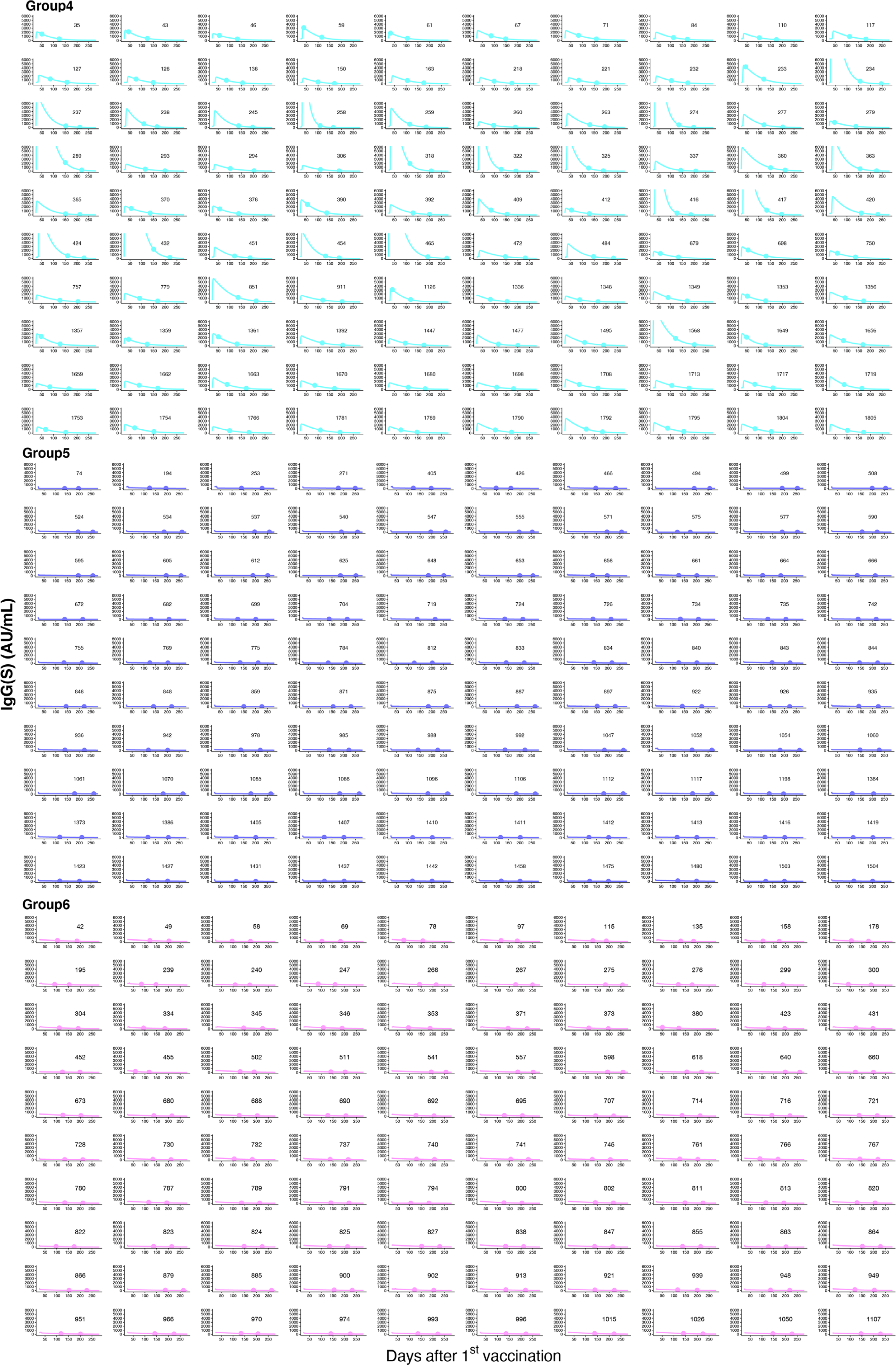
Antibody titer trajectory for individual participants stratified into different groups: The estimated antibody titer for each individual participant (solid lines) along with the observed data (closed dots) are depicted using the best-fit parameter estimates. G1, G2, G3, G4, G5, and G6 are shown in yellow, gray, green, light blue, blue, and pink, respectively. In each group, the curve of 100 was randomly selected for visualization because of the large number.

**Extended Data Table 1.**
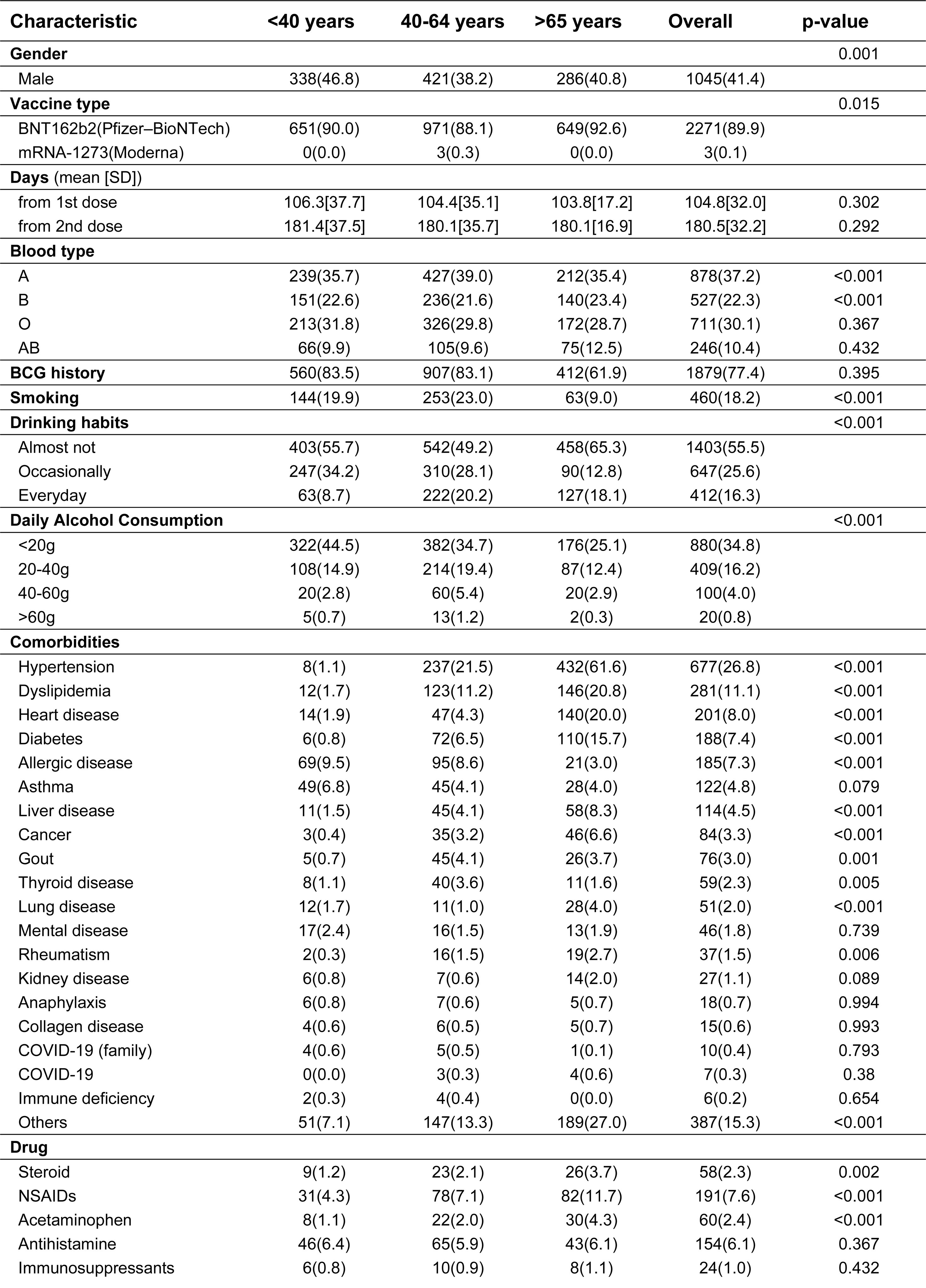
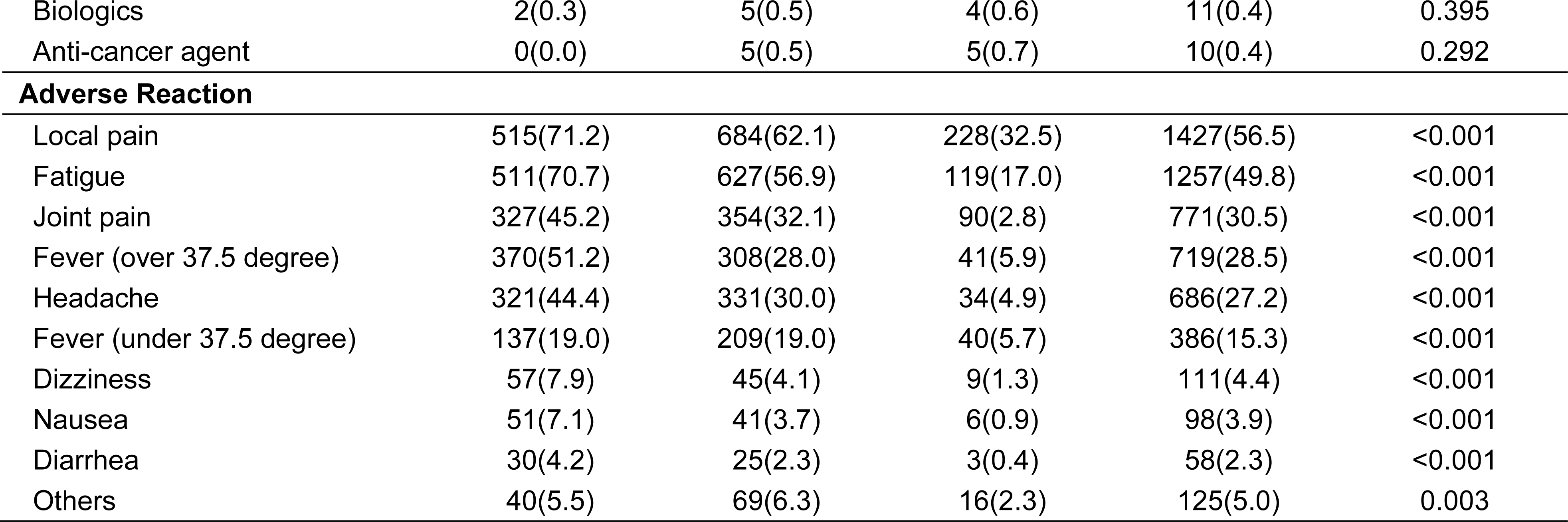
Basic demographics for the Fukushima vaccination cohort.

**Supplementary Table 1.** Details of query words and results of the literature review.

| # | Query Word | Results |
| --- | --- | --- |
| 1 | "SARS-CoV-2" [tiab] OR "severe coronavirus disease 2019" [tiab] OR "COVID-19" [tiab] OR "severe acute respiratory syndrome coronavirus 2" [tiab] OR "coronavirus disease 2019" [tiab] OR "COVID-19*" [Mesh] OR "SARS-CoV-2" | 235,156 |
| 2 | "vaccination*" [tiab] OR "vaccine*" [tiab] OR "NVX-CoV2373" [tiab] OR "CVnCoV" [tiab] OR "Ad26COV2S" [tiab] OR "BNT162 Vaccine*" [Mesh] OR "ChAdOx1 nCoV-19" [Mesh] OR "COVID-19 Vaccines*" [Mesh] OR "mRNA Vaccines" [Mesh] OR "RNA, Viral" [Mesh] OR "Vaccination*" [Mesh] OR "Vaccine Efficacy*" [Mesh] OR "Vaccines*" [Mesh] OR "2019-nCoV Vaccine mRNA-1273*" [Mesh] OR "Ad26COVS1" [Mesh] | 500,257 |
| 3 | "Antibody level*" [tiab] OR "antibody response*" [tiab] OR "Antibody titer*" [tiab] OR "Humoral and Cellular Response*" [tiab] OR "humoral immune response*" [tiab] OR "humoral response*" [tiab] OR "immune response*" [tiab] OR "immunization" [tiab] OR "immunogenicity" [tiab] OR "Antibodies, Viral" [Mesh] OR "Antibody Formation" [Mesh] OR "Immunity, Humoral*" [Mesh] OR "Immunogenicity, Vaccine*" [Mesh] | 557,862 |
| 4 | "Second dose" [tiab] OR "second SARS-CoV-2 vaccine" [tiab] OR "2nd dose" [tiab] OR "second vaccination" [tiab] OR "two doses" [tiab] OR "two sequential doses" [tiab] OR "Two Vaccine Doses" [tiab] OR "two-dose" [tiab] OR "booster doses" [tiab] OR "second vaccine" [tiab] OR "Immunization, Secondary*" [Mesh] | 30,432 |
| 5 | "Cohort" [tiab] OR "Cross-Sectional" [tiab] OR "Follow-Up Studies" [tiab] OR "Prospective Studies" [tiab] OR "Cohort Studies" [Mesh] OR "Cross-Sectional Studies" [Mesh] OR "Follow-Up Studies" [Mesh] OR "Prospective Studies" [Mesh] OR "Observational Study*" [tiab] | 3,119,085 |
| 6 | 1 AND 2 | 32,087 |
| 7 | 1 AND 2 AND 3 | 7,561 |
| 8 | 1 AND 2 AND 3 AND 4 | 1,311 |
| 9 | 1 AND 2 AND 3 AND 4 AND 5 | 440 |

**Supplementary Table 2.** Characteristics of the previously reported cohorts.

| # | Cohort Size | Study Period | Country | Vaccine Type | Population Characteristics | Interview Sheet | Times of Sampling | Sample 1 | Sample 2 | Sample final | Number of Participants Included in All Sampling | Samples of Neutralizing Antibody | Sample1 of Neutralizing Antibody | Reference |
| --- | --- | --- | --- | --- | --- | --- | --- | --- | --- | --- | --- | --- | --- | --- |
| 0 | 2526 | 270 | Japan | BNT162b2 (Pfizer–BioNTech) or mRNA-1273 (Moderna) | Community | Including | 2 | 2526 | 2443 | 2443 | 2443 | Including | 2526 | This study |
| 1 | 3,991 | 200 | Israel | BNT162b2 (Pfizer–BioNTech) | HCWs | Including | 7 | 3991 | 2690 | 1370 | 693 | Including | 681 | 1 |
| 2 | 2,591 | 180 | Italy | BNT162b2 (Pfizer–BioNTech) | Community | Not Including | 4 | 1725 | 1641 | 2591 | 1215 | Not Including | NA | 2 |
| 3 | 1,935 | 90 | Italy | BNT162b2 (Pfizer–BioNTech) | HCWs | Not Including | 3 | 1935 | NA | NA | NA | Including | 1935 | 3 |
| 4 | 1,506 | 90 | Israel | BNT162b2 (Pfizer–BioNTech) | HCWs | Including | 2 | 1506 | 1209 | 1209 | 1194 | Not Including | NA | 4 |
| 5 | 1,487 | 41 | Israel | BNT162b2 (Pfizer–BioNTech) | HCWs | Including | 5 | 334 | 375 | 1487 | 22 | Including | 46 | 5 |
| 6 | 1,012 | 102 | Turkey | CoronaVac (Sinovac) | HCWs | Including | 2 | 1012 | 836 | 836 | 836 | Not Including | NA | 6 |

**Supplementary Table 3.**
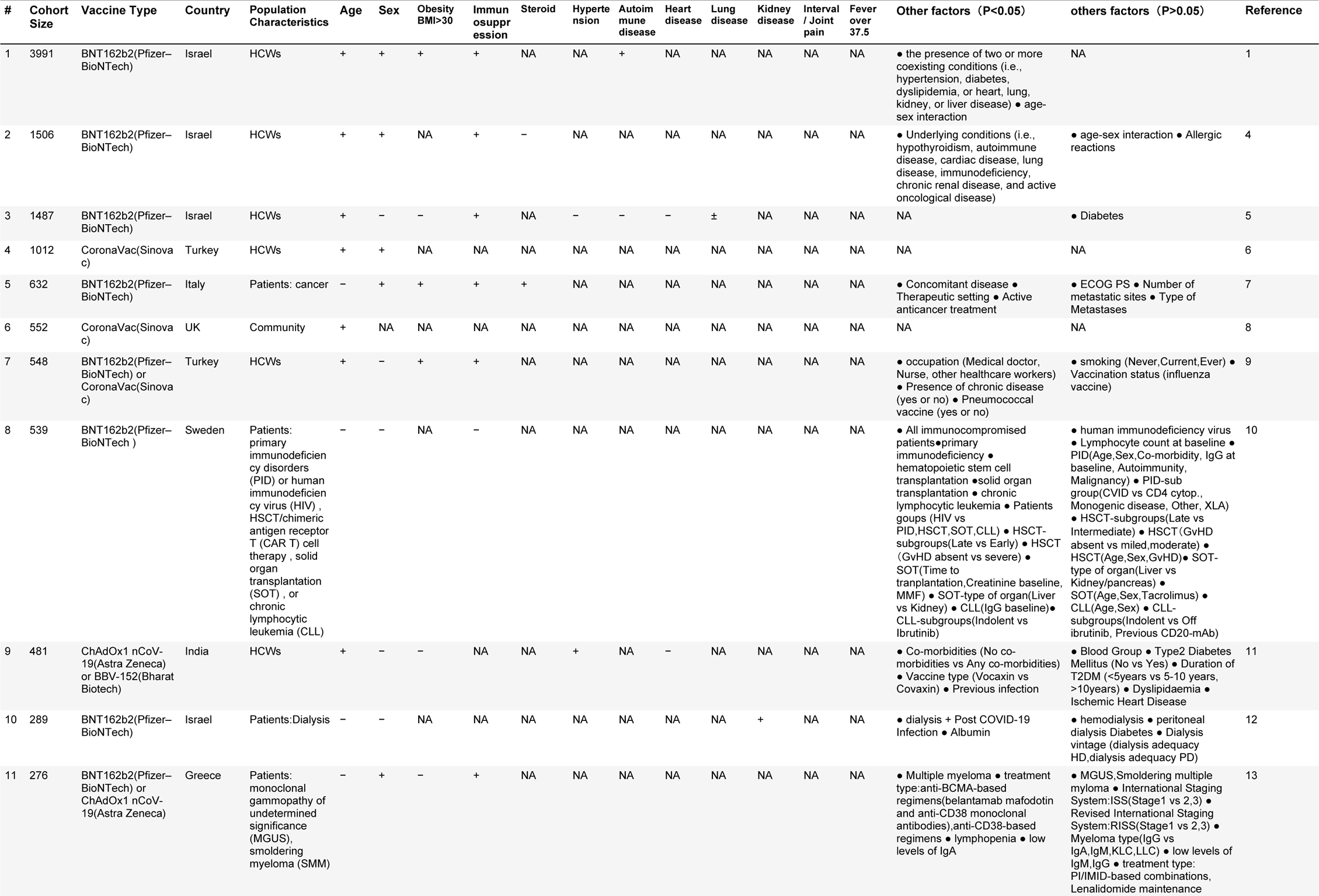
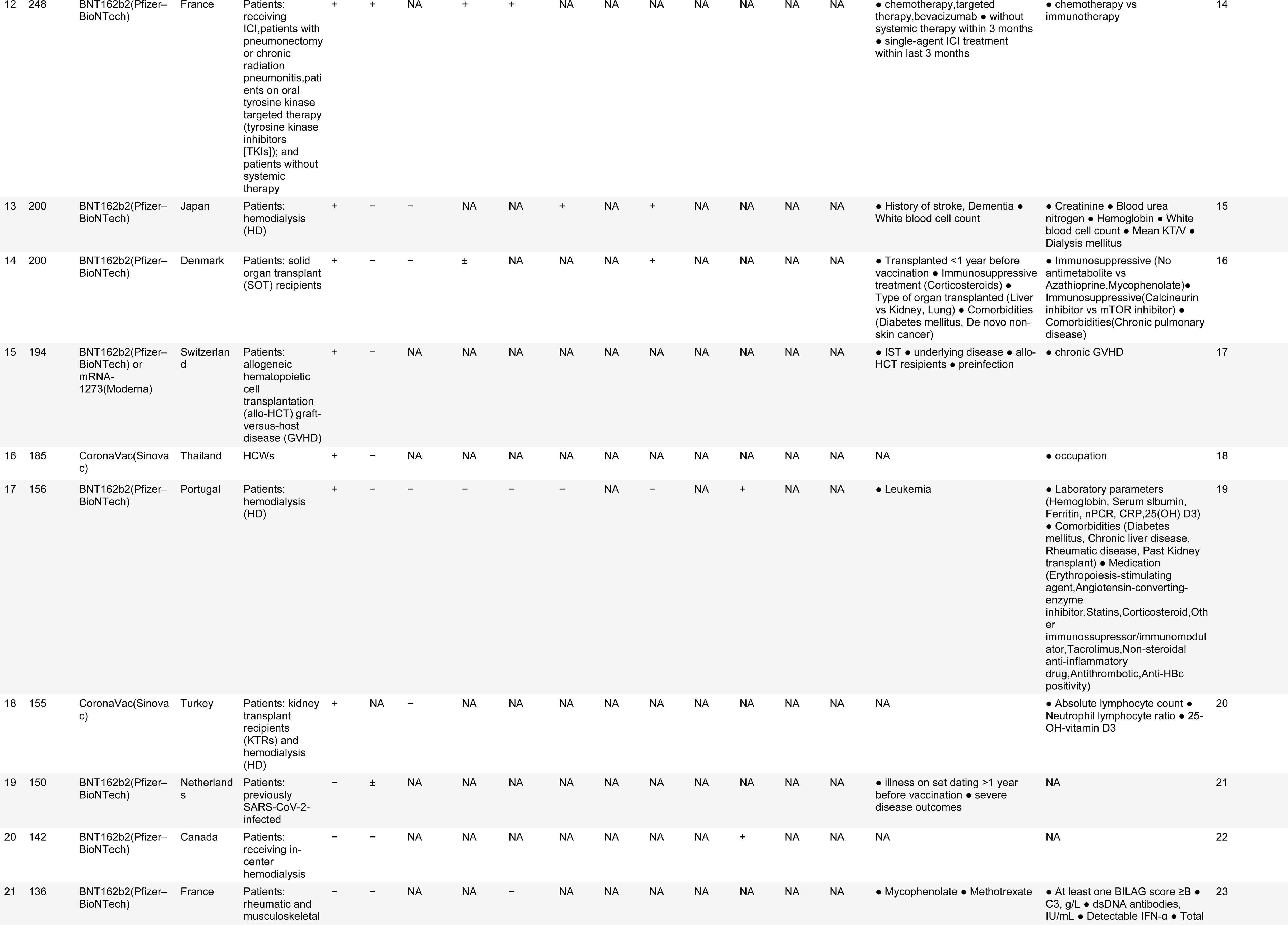
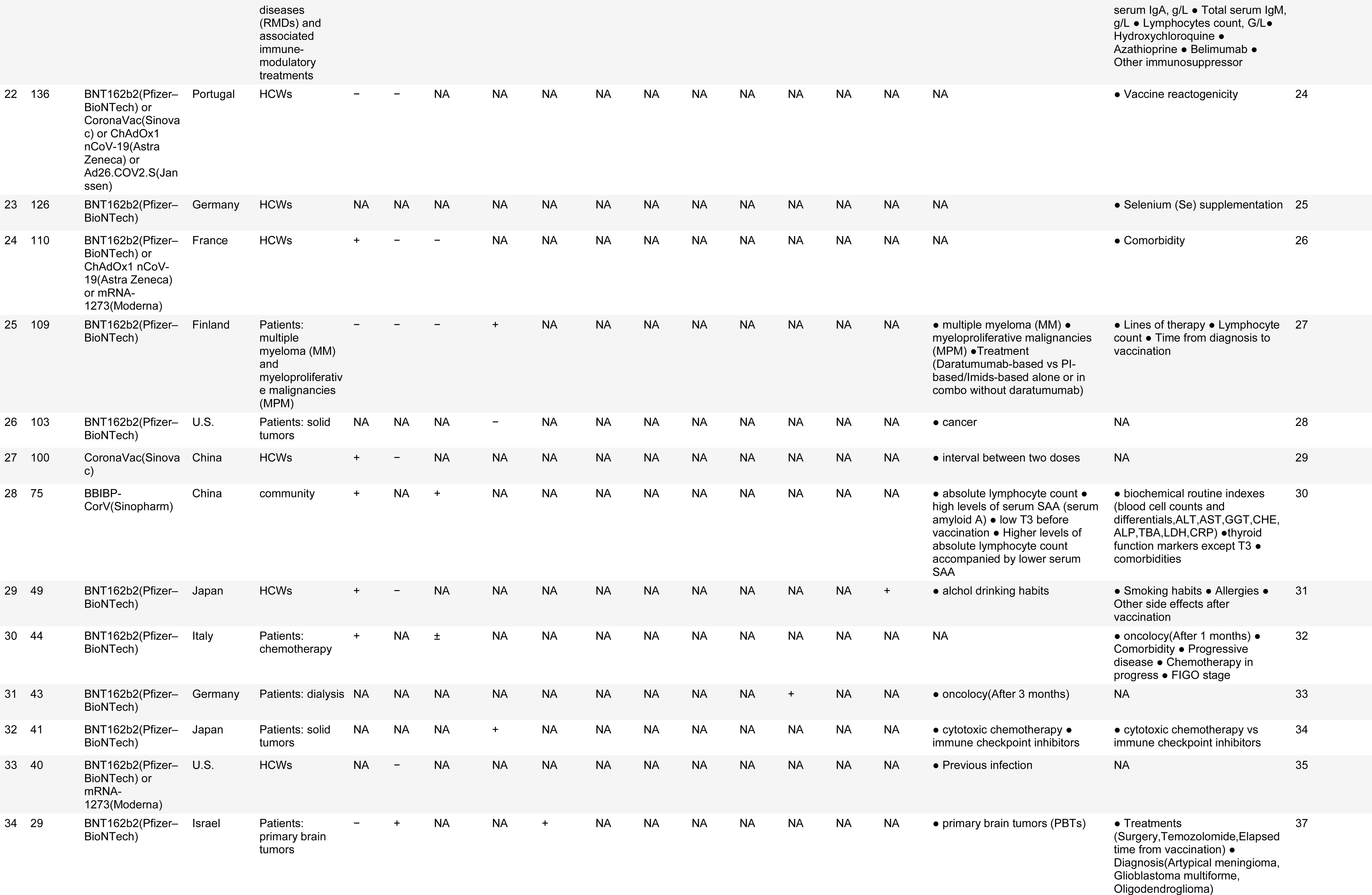
Characteristics of the vulnerable population.

| # | Cohort Size | Vaccine Type | Country | Population Characteristics | Age | Sex | Obesity BMI>30 | Immunosuppression | Steroid | Hypertension | Autoimmune disease | Heart disease | Lung disease | Kidney disease | Interval / Joint pain | Fever over 37.5 | Other factors (P<0.05) | others factors (P>0.05) | Reference |
| --- | --- | --- | --- | --- | --- | --- | --- | --- | --- | --- | --- | --- | --- | --- | --- | --- | --- | --- | --- |
| 1 | 3991 | BNT162b2(Pfizer–BioNTech) | Israel | HCWs | + | + | + | + | NA | NA | + | NA | NA | NA | NA | NA | the presence of two or more coexisting conditions (i.e., hypertension, diabetes, dyslipidemia, or heart, lung, kidney, or liver disease)<br>age-sex interaction | NA | 1 |
| 2 | 1506 | BNT162b2(Pfizer–BioNTech) | Israel | HCWs | + | + | NA | + | – | NA | NA | NA | NA | NA | NA | NA | Underlying conditions (i.e., hypothyroidism, autoimmune disease, cardiac disease, lung disease, immunodeficiency, chronic renal disease, and active oncological disease) | age-sex interaction<br>Allergic reactions | 4 |
| 3 | 1487 | BNT162b2(Pfizer–BioNTech) | Israel | HCWs | + | – | – | + | NA | – | – | – | ± | NA | NA | NA | NA | Diabetes | 5 |
| 4 | 1012 | CoronaVac(Sinovac) | Turkey | HCWs | + | + | NA | NA | NA | NA | NA | NA | NA | NA | NA | NA | NA | NA | 6 |
| 5 | 632 | BNT162b2(Pfizer–BioNTech) | Italy | Patients: cancer | – | + | + | + | + | NA | NA | NA | NA | NA | NA | NA | Concomitant disease<br>Therapeutic setting<br>Active anticancer treatment | ECOG PS<br>Number of metastatic sites<br>Type of Metastases | 7 |
| 6 | 552 | CoronaVac(Sinovac) | UK | Community | + | NA | NA | NA | NA | NA | NA | NA | NA | NA | NA | NA | NA | NA | 8 |
| 7 | 548 | BNT162b2(Pfizer–BioNTech) or CoronaVac(Sinovac) | Turkey | HCWs | + | – | + | + | NA | NA | NA | NA | NA | NA | NA | NA | occupation (Medical doctor, Nurse, other healthcare workers)<br>Presence of chronic disease (yes or no)<br>Pneumococcal vaccine (yes or no) | smoking (Never,Current,Ever)<br>Vaccination status (influenza vaccine) | 9 |
| 8 | 539 | BNT162b2(Pfizer–BioNTech) | Sweden | Patients: primary immunodeficiency disorders (PID) or human immunodeficiency virus (HIV), HSCT/chimeric antigen receptor T (CAR T) cell therapy, solid organ transplantation (SOT), or chronic lymphocytic leukemia (CLL) | – | – | NA | – | NA | NA | NA | NA | NA | NA | NA | NA | All immunocompromised patients<br>primary immunodeficiency<br>hematopoietic stem cell transplantation<br>solid organ transplantation<br>chronic lymphocytic leukemia<br>Patients groups (HIV vs PID, HSCT, SOT, CLL)<br>HSCT-subgroups(Late vs Early)<br>HSCT (GvHD absent vs severe)<br>SOT(Time to transplantation, Creatinine baseline, MMF)<br>SOT-type of organ(Liver vs Kidney)<br>CLL(IgG baseline)<br>CLL-subgroups(Indolent vs Ibrutinib) | human immunodeficiency virus<br>Lymphocyte count at baseline<br>PID(Age,Sex,Co-morbidity, IgG at baseline, Autoimmunity, Malignancy)<br>PID-sub group(CVID vs CD4 cytop., Monogenic disease, Other, XLA)<br>HSCT-subgroups(Late vs Intermediate)<br>HSCT (GvHD absent vs mild, moderate)<br>HSCT(Age,Sex,GvHD)<br>SOT-type of organ(Liver vs Kidney/pancreas)<br>SOT(Age,Sex,Tacrolimus)<br>CLL(Age,Sex)<br>CLL-subgroups(Indolent vs Off ibritinib, Previous CD20-mAb) | 10 |
| 9 | 481 | ChAdOx1 nCoV-19(Astra Zeneca) or BBV-152(Bharat Biotech) | India | HCWs | + | – | – | NA | NA | + | NA | – | NA | NA | NA | NA | Co-morbidities (No co-morbidities vs Any co-morbidities)<br>Vaccine type (Vocaxin vs Covaxin)<br>Previous infection | Blood Group<br>Type2 Diabetes Mellitus (No vs Yes)<br>Duration of T2DM (<5years vs 5-10 years, >10years)<br>Dyslipidaemia<br>Ischemic Heart Disease | 11 |
| 10 | 289 | BNT162b2(Pfizer–BioNTech) | Israel | Patients:Dialysis | – | – | NA | NA | NA | NA | NA | NA | NA | + | NA | NA | dialysis + Post COVID-19 Infection<br>Albumin | hemodialysis<br>peritoneal dialysis<br>Diabetes<br>Dialysis vintage (dialysis adequacy HD,dialysis adequacy PD) | 12 |
| 11 | 276 | BNT162b2(Pfizer–BioNTech) or ChAdOx1 nCoV-19(Astra Zeneca) | Greece | Patients: monoclonal gammopathy of undetermined significance (MGUS), smoldering myeloma (SMM) | – | + | – | + | NA | NA | NA | NA | NA | NA | NA | NA | Multiple myeloma<br>treatment type:anti-BCMA-based regimens(belantamab mafodotin and anti-CD38 monoclonal antibodies),anti-CD38-based regimens<br>lymphopenia<br>low levels of IgA | MGUS,Smoldering multiple myeloma<br>International Staging System:ISS(Stage1 vs 2,3)<br>Revised International Staging System:RISS(Stage1 vs 2,3)<br>Myeloma type(IgG vs IgA,IgM,KLC,LLC)<br>low levels of IgM,IgG<br>treatment type: PI/IMiD-based combinations, Lenalidomide maintenance | 13 |
| 12 | 248 | BNT162b2(Pfizer–BioNTech) | France | Patients: receiving ICI, patients with pneumonectomy or chronic radiation pneumonitis, patients on oral tyrosine kinase targeted therapy (tyrosine kinase inhibitors [TKIs]); and patients without systemic therapy | + | + | NA | + | + | NA | NA | NA | NA | NA | NA | NA | chemotherapy, targeted therapy, bevacizumab<br>without systemic therapy within 3 months<br>single-agent ICI treatment within last 3 months | chemotherapy vs immunotherapy | 14 |
| 13 | 200 | BNT162b2(Pfizer–BioNTech) | Japan | Patients: hemodialysis (HD) | + | – | – | NA | NA | + | NA | + | NA | NA | NA | NA | History of stroke, Dementia<br>White blood cell count | Creatinine<br>Blood urea nitrogen<br>Hemoglobin<br>White blood cell count<br>Mean KT/V<br>Dialysis mellitus | 15 |
| 14 | 200 | BNT162b2(Pfizer–BioNTech) | Denmark | Patients: solid organ transplant (SOT) recipients | + | – | – | ± | NA | NA | NA | + | NA | NA | NA | NA | Transplanted <1 year before vaccination<br>Immunosuppressive treatment (Corticosteroids)<br>Type of organ transplanted (Liver vs Kidney, Lung)<br>Comorbidities (Diabetes mellitus, De novo non-skin cancer) | Immunosuppressive (No antimetabolite vs Azathioprine, Mycophenolate)<br>Immunosuppressive (Calcineurin inhibitor vs mTOR inhibitor)<br>Comorbidities (Chronic pulmonary disease) | 16 |
| 15 | 194 | BNT162b2(Pfizer–BioNTech) or mRNA-1273 (Moderna) | Switzerland | Patients: allogeneic hematopoietic cell transplantation (allo-HCT) graft-versus-host disease (GVHD) | + | – | NA | NA | NA | NA | NA | NA | NA | NA | NA | NA | IST<br>underlying disease<br>allo-HCT recipients<br>preinfection | chronic GVHD | 17 |
| 16 | 185 | CoronaVac (Sinovac) | Thailand | HCWs | + | – | NA | NA | NA | NA | NA | NA | NA | NA | NA | NA | NA | occupation | 18 |
| 17 | 156 | BNT162b2(Pfizer–BioNTech) | Portugal | Patients: hemodialysis (HD) | + | – | – | – | – | – | NA | – | NA | + | NA | NA | Leukemia | Laboratory parameters (Hemoglobin, Serum albumin, Ferritin, nPCR, CRP, 25(OH) D3)<br>Comorbidities (Diabetes mellitus, Chronic liver disease, Rheumatic disease, Past Kidney transplant)<br>Medication (Erythropoiesis-stimulating agent, Angiotensin-converting enzyme inhibitor, Statins, Corticosteroid, Other immunosuppressor/immunomodulator, Tacrolimus, Non-steroidal anti-inflammatory drug, Antithrombotic, Anti-HBc positivity) | 19 |
| 18 | 155 | CoronaVac (Sinovac) | Turkey | Patients: kidney transplant recipients (KTRs) and hemodialysis (HD) | + | NA | – | NA | NA | NA | NA | NA | NA | NA | NA | NA | NA | Absolute lymphocyte count<br>Neutrophil lymphocyte ratio<br>25-OH-vitamin D3 | 20 |
| 19 | 150 | BNT162b2(Pfizer–BioNTech) | Netherlands | Patients: previously SARS-CoV-2-infected | – | ± | NA | NA | NA | NA | NA | NA | NA | NA | NA | NA | illness on set dating >1 year before vaccination<br>severe disease outcomes | NA | 21 |
| 20 | 142 | BNT162b2(Pfizer–BioNTech) | Canada | Patients: receiving in-center hemodialysis | – | – | NA | NA | NA | NA | NA | NA | NA | + | NA | NA | NA | NA | 22 |
| 21 | 136 | BNT162b2(Pfizer–BioNTech) | France | Patients: rheumatic and musculoskeletal | – | – | NA | NA | – | NA | NA | NA | NA | NA | NA | NA | Mycophenolate<br>Methotrexate | At least one BILAG score ≥B<br>C3, g/L<br>dsDNA antibodies, IU/mL<br>Detectable IFN-α<br>Total | 23 |

|  |  |  |  | diseases (RMDs) and associated immunomodulatory treatments |  |  |  |  |  |  |  |  |  |  |  |  |  | serum IgA, g/L • Total serum IgM, g/L • Lymphocytes count, G/L • Hydroxychloroquine • Azathioprine • Belimumab • Other immunosuppressor |  |
| --- | --- | --- | --- | --- | --- | --- | --- | --- | --- | --- | --- | --- | --- | --- | --- | --- | --- | --- | --- |
| 22 | 136 | BNT162b2(Pfizer–BioNTech) or CoronaVac(Sinovac) or ChAdOx1 nCoV-19(AstraZeneca) or Ad26.COV2.S(Janssen) | Portugal | HCWs | – | – | NA | NA | NA | NA | NA | NA | NA | NA | NA | NA | NA | • Vaccine reactogenicity | 24 |
| 23 | 126 | BNT162b2(Pfizer–BioNTech) | Germany | HCWs | NA | NA | NA | NA | NA | NA | NA | NA | NA | NA | NA | NA | NA | • Selenium (Se) supplementation | 25 |
| 24 | 110 | BNT162b2(Pfizer–BioNTech) or ChAdOx1 nCoV-19(AstraZeneca) or mRNA-1273(Moderna) | France | HCWs | + | – | – | NA | NA | NA | NA | NA | NA | NA | NA | NA | NA | • Comorbidity | 26 |
| 25 | 109 | BNT162b2(Pfizer–BioNTech) | Finland | Patients: multiple myeloma (MM) and myeloproliferative malignancies (MPM) | – | – | – | + | NA | NA | NA | NA | NA | NA | NA | NA | • multiple myeloma (MM) • myeloproliferative malignancies (MPM) • Treatment (Daratumumab-based vs PI-based/Imids-based alone or in combo without daratumumab) | • Lines of therapy • Lymphocyte count • Time from diagnosis to vaccination | 27 |
| 26 | 103 | BNT162b2(Pfizer–BioNTech) | U.S. | Patients: solid tumors | NA | NA | NA | – | NA | NA | NA | NA | NA | NA | NA | NA | • cancer | NA | 28 |
| 27 | 100 | CoronaVac(Sinovac) | China | HCWs | + | – | NA | NA | NA | NA | NA | NA | NA | NA | NA | NA | • interval between two doses | NA | 29 |
| 28 | 75 | BBIBP-CorV(Sinopharm) | China | community | + | NA | + | NA | NA | NA | NA | NA | NA | NA | NA | NA | • absolute lymphocyte count • high levels of serum SAA (serum amyloid A) • low T3 before vaccination • Higher levels of absolute lymphocyte count accompanied by lower serum SAA | • biochemical routine indexes (blood cell counts and differentials,ALT,AST,GGT,CHE,ALP,TBA,LDH,CRP) •thyroid function markers except T3 • comorbidities | 30 |
| 29 | 49 | BNT162b2(Pfizer–BioNTech) | Japan | HCWs | + | – | NA | NA | NA | NA | NA | NA | NA | NA | NA | + | • alchol drinking habits | • Smoking habits • Allergies • Other side effects after vaccination | 31 |
| 30 | 44 | BNT162b2(Pfizer–BioNTech) | Italy | Patients: chemotherapy | + | NA | ± | NA | NA | NA | NA | NA | NA | NA | NA | NA | NA | • oncology(After 1 months) • Comorbidity • Progressive disease • Chemotherapy in progress • FIGO stage | 32 |
| 31 | 43 | BNT162b2(Pfizer–BioNTech) | Germany | Patients: dialysis | NA | NA | NA | NA | NA | NA | NA | NA | NA | + | NA | NA | • oncology(After 3 months) | NA | 33 |
| 32 | 41 | BNT162b2(Pfizer–BioNTech) | Japan | Patients: solid tumors | NA | NA | NA | + | NA | NA | NA | NA | NA | NA | NA | NA | • cytotoxic chemotherapy • immune checkpoint inhibitors | • cytotoxic chemotherapy vs immune checkpoint inhibitors | 34 |
| 33 | 40 | BNT162b2(Pfizer–BioNTech) or mRNA-1273(Moderna) | U.S. | HCWs | NA | – | NA | NA | NA | NA | NA | NA | NA | NA | NA | NA | • Previous infection | NA | 35 |
| 34 | 29 | BNT162b2(Pfizer–BioNTech) | Israel | Patients: primary brain tumors | – | + | NA | NA | + | NA | NA | NA | NA | NA | NA | NA | • primary brain tumors (PBTs) | • Treatments (Surgery,Temozolomide,Elapsed time from vaccination) • Diagnosis(Artypical meningioma, Glioblastoma multiforme, Oligodendrogloma) | 37 |

**Supplementary Table 4.** Estimated fixed and individual parameters for 12 health care workers.

| Parameter or variable | Decay rate of antibody-secreting cells | Maximum *de novo* production of antibody by 1<sup>st</sup> vaccination | Delay of antibody-secreting cells induction after 1<sup>st</sup> vaccination. | Steepness at which induction increases with increasing the amount of mRNA | Amount of mRNA satisfying $P_i/2$ | Maximum *de novo* production of antibody by 2<sup>nd</sup> vaccination | Delay of antibody-secreting cells induction after 2<sup>nd</sup> vaccination. |
| --- | --- | --- | --- | --- | --- | --- | --- |
| Symbol | $\mu$ | $H_1$ | $\eta_1$ | $m$ | $K$ | $H_2$ | $\eta_2$ |
| Unit | day<sup>-1</sup> | AU/mL | day | --- | $\mu\text{g}/0.5\text{mL}$ | AU/mL | day |
| **Individual estimated parameters for $S_1$ to $S_{12}$** |  |  |  |  |  |  |  |
| $S_1$ | 0.885346 | 650.518 | 12.5269 | 0.0373144 | 33900 | 5227.34 | 4.21433 |
| $S_2$ | 0.885346 | 760.114 | 12.5109 | 0.028673 | 33900 | 5035.79 | 4.23423 |
| $S_3$ | 0.885346 | 2375.11 | 12.5096 | 0.0223842 | 33900 | 7562.82 | 3.85759 |
| $S_4$ | 0.885346 | 790.745 | 12.4927 | 0.0611514 | 33900 | 7304.66 | 3.95306 |
| $S_5$ | 0.885346 | 942.686 | 12.5169 | 0.0250174 | 33900 | 5638.12 | 3.91276 |
| $S_6$ | 0.885346 | 448.87 | 12.5243 | 0.0851651 | 33900 | 3828.1 | 4.24752 |
| $S_7$ | 0.885346 | 1848.01 | 12.5471 | 0.0272096 | 33900 | 9073.94 | 4.2513 |
| $S_8$ | 0.885346 | 567.839 | 12.5409 | 0.0523284 | 33900 | 4673.2 | 4.71178 |
| $S_9$ | 0.885346 | 1835.9 | 12.5428 | 0.0365247 | 33900 | 7517.48 | 4.59114 |
| $S_{10}$ | 0.885346 | 1201.77 | 12.5109 | 0.0625655 | 33900 | 5916.28 | 4.17894 |
| $S_{11}$ | 0.885346 | 736.229 | 12.5086 | 0.0700219 | 33900 | 4118.67 | 4.54006 |
| $S_{12}$ | 0.885346 | 1369.65 | 12.5409 | 0.0488345 | 33900 | 8226.08 | 4.23969 |
| **Population estimated parameters** |  |  |  |  |  |  |  |
| --- | 0.885346 | 975.297 | 12.52 | 0.0437653 | 33900 | 6035.142 | 4.25 |

